# A variant of the autophagic receptor NDP52 counteracts phospho-TAU accumulation and emerges as a protective factor for Alzheimer Disease

**DOI:** 10.1101/2024.08.13.24311780

**Authors:** Anna Mattioni, Claudia Carsetti, Krenare Bruqi, Valerio Caputo, Valentina Cianfanelli, Maria Giulia Bacalini, Mariella Casa, Carlo Gabelli, Emiliano Giardina, Gianluca Cestra, Flavie Strappazzon

**Affiliations:** IRCCS Fondazione Santa Lucia, Via Ardeatina 306/354, 00179, Rome, Italy; Institute of Molecular Biology and Pathology (IBPM), National Research Council (CNR), c/o University of Rome Sapienza, Piazzale Aldo Moro 5, 00185 Rome, Italy; University of Rome Sapienza, Department of Biology and Biotechnology, Piazzale Aldo Moro 5, 00185, Rome, Italy; Univ Lyon, Univ Lyon 1, CNRS, INSERM, Physiopathologie et Génétique du Neurone et du muscle, UMR5261, U1315, Institut Neuromyogène, 8 avenue Rockefeller, 69008 Lyon, France; Department of Clinical Medicine, Life, Health & Environmental Sciences-MESVA, University of L’Aquila; Department of Science, University "ROMA TRE", Viale Guglielmo Marconi 446, 00146, Rome, Italy; Department of Woman and Child Health and Public Health, Gynecologic Oncology Unit, Fondazione Policlinico Universitario A. Gemelli IRCCS, Largo Agostino Gemelli 8, 00168, Rome, Italy; IRCCS Istituto delle Scienze Neurologiche di Bologna, Via Altura 3, 40139 Bologna, Italy; Clinical Center for the Aging Brain, Dept of Medicine, University of Padua, Via Giustiniani 2, 35128 Padova, Italy; Department of Biomedicine and Prevention, Tor Vergata University, Viale Montpellier 1, 00133, Rome, Italy

**Keywords:** Alzheimer disease, AT8, AT100, NDP52/CALCOCO2, PP2A, pSer396TAU, Selective autophagy, Tauopathy

## Abstract

Selective elimination of early pathological TAU species may be a promising therapeutic strategy to reduce TAU accumulation that contributes to neurodegeneration and hallmarks Alzheimer disease (AD). By performing a genetic analysis of a cohort of 435 patients with AD, we defined the NDP52^GE^ variant (rs550510) of the autophagic receptor NDP52 (also known as CALCOCO2) as a protective factor for AD. We provide evidence that in *in vitro* systems and in a *Drosophila melanogaster* model of TAU-induced AD, NDP52 reduces the accumulation of pathological forms of TAU through the autophagic process and rescues typical neurodegenerative phenotypes induced by hTAU-toxicity. More importantly, we showed that the NDP52^GE^ variant is much more effective in this respect than NDP52^WT^. Mechanistically, we showed that NDP52 directly binds pathological phospho-TAU, and that NDP52^WT^ and NDP52^GE^ bind them with comparable efficiency. On the contrary, we showed that NDP52^GE^ binds the autophagic machinery (LC3C and LC3B) more efficiently than NDP52^WT^. We also showed for the first time that NDP52 is a direct target of protein phosphatase 2A (PP2A) *in vitro*, opening the way to the possibility that this phosphatase may fine-tune the autophagic function of NDP52 in AD. Finally, we found a positive correlation between the worldwide distribution of the allele encoding NDP52^GE^ and the incidence or prevalence of AD. Overall, our work highlights the variant NDP52^GE^ as a resilience factor in AD that shows a robust effectiveness to drive pathological TAU degradation.

## INTRODUCTION

Alzheimer disease (AD) is the most prevalent progressive neurological disorder affecting aged people worldwide [1,2]. In the brain, Amyloid-β (Aβ) plaques and TAU neurofibrillary tangles pathologically characterize AD and are associated with loss of synapses and neurons, which result in dementia [3]. To date, no effective disease-modifying therapies are available, claiming for a deeper understanding of the molecular mechanisms leading to neuronal death and for the identification of new potential therapeutic targets [4,5]. Accumulating evidence suggests that TAU pathology – which consists in TAU mis-localization, oligomerization, changes in solubility, and aggregation in filaments and neurofibrillary tangles - correlates with the degree of dementia better than Aβ deposition [6–8]. Furthermore, the early phases of TAU pathology, mainly triggered by the hyperphosphorylation of TAU, appear to correlate better with neuronal toxicity than the highly ordered filaments typical of the final phases of the disease [9–11]. The clearance of these early pathological TAU species, targeting in particular hyperphosphorylated TAU, could be therefore a promising strategy for therapeutic interventions.

NDP52 (Nuclear Dot Protein 52, also known as CALCOCO2) is an autophagic receptor that has been shown to facilitate pathological phospho-TAU (pTAU) clearance via selective autophagy [12]. Mechanistically, as the other autophagic receptors, NDP52 physically links specific cargoes committed to lysosomal degradation, with the autophagy core machinery responsible for the formation of the cup-shaped membrane called phagophore. This initial step triggers the signaling cascade that allows cargo enwrapment by forming a double-membrane vesicle - the autophagosome - which then fuses with lysosomes for content degradation [13]. The interaction between the cLIR (non-canonical LC3 interacting region) motif of NDP52 and LC3C, an ATG8 family member anchored to the nascent phagophore, ensures the efficient engulfment of the cargo selected by NDP52 in the autophagosome. In addition, the binding of a distinct LIR motif of NDP52 with other ATG8 family members (LC3A/B and GABARAPs, but not LC3C), and the interaction with the motor protein MYOSIN VI, allow NDP52 to promote the autophagosome maturation, ensuring efficient cargoes degradation [13,14].

In mouse and rat cortical neurons, silencing or overexpression of NDP52 resulted in increased or reduced levels of pathological phospho-TAU (pTAU), respectively. Moreover, in both AD mouse brain and AD human cerebral cortex, NDP52 interacts with pathological pTAU, and the SKICH (skeletal muscle and kidney-enriched inositol phosphatase (SKIP) carboxyl homology) domain of NDP52 binds TAU *in vitro* [12,15]. Altogether, these data strongly suggest that NDP52 could prevent TAU aggregation by clearing pathological pTAU, therefore slowing the progression of AD.

In 2021, we characterized a variant of NDP52, hereafter referred to as NDP52^GE^ (c.491G>A, rs550510, p.G140E), in which the G140E substitution enhances the ability of NDP52 to interact with LC3C. Indeed, this amino acidic position is very close to the cLIR motif and the presence of a glutamic acid residue (instead of a glycine) appears to induce a conformational change that favours the interaction with LC3C. We also showed that, compared to its wild-type form (NDP52^WT^), NDP52^GE^ promotes a stronger degradation of damaged mitochondria – a well-known cargo of NDP52 [16].

Here, we explored the hypothesis that NDP52^GE^, by promoting a greater autophagic degradation of pathological pTAU, could counter its accumulation better than NDP52^WT^, delaying the onset and/or the progression of AD.

## RESULTS

### The variant NDP52 G140E is associated to and protective in AD

To explore a possible protective role of the NDP52^GE^ variant in Alzheimer Disease (AD), we performed a genetic association case-control analysis on a cohort of 434 AD patients and 1000 control subjects. The results, reported in Table 1, showed that the frequency of the allele A (c.491G>A, rs550510, p.G140E), coding for NDP52^GE^, was lower in the AD group (cases) compared to the control group. The biostatistical analysis indicated that the association with the disease is statistically significant (*p* = 0.0042), and that the variant allele A is protective in AD (Odds Ratio, OR A= 0.72, 95%CI: 0.58-0.90) (Table 1). Similarly, the frequency of the heterozygous genotype (GA) and that of the homozygous genotype for the variant allele (AA) were lower in the AD group compared to the control group. Also in this case, the biostatistical analysis confirmed that the associations with the disease are statistically significant (*p* = 0,017), with the homozygous genotype for the variant allele (AA) being protective in AD (OR AA= 0.52, 95% CI: 0.26 - 1.05). Taken together, these results indicate that the NDP52^GE^ variant could be a protective factor in AD.

**Table 1.**
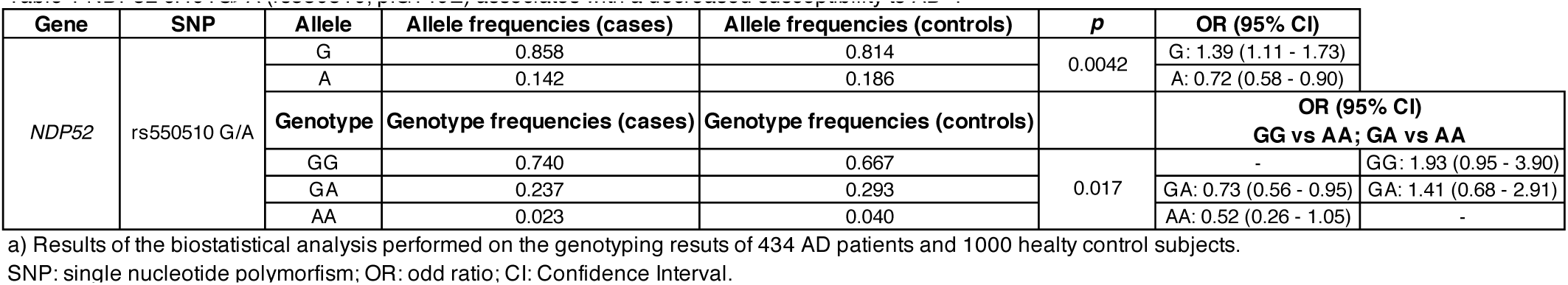
NDP52 c.491G>A (rs550510, p.G140E) associates with a decreased susceptibility to AD^a)^.

### In the human SH-SY5Y neuroblastoma cell line, NDP52^GE^ variant binds LC3C more efficiently than NDP52^WT^

To characterize the protective role of NDP52^GE^ in AD and its involvement in the autophagic process, we firstly compared the binding of NDP52^WT^ and NDP52^GE^ to the different members of the ATG8 family. We performed co-immunoprecipitation assays from human neuroblastoma SH-SY5Y cells that overexpressed each ATG8 family member with NDP52^WT^ or NDP52^GE^ (the NDP52 constructs were schematically represented in Figure 2A). The results showed that both NDP52 proteins strongly interact with LC3C, while their binding to LC3B and LC3A was weak (FigS1B) and those to GABARAPs even lower (FigS1A). Moreover, NDP52^GE^ binding to GABARAPs showed an efficiency comparable to that of NDP52^WT^ (figure S1A), while binding to LC3A tended to increase compared to NDP52^WT^ without however reaching statistical significance. On the contrary, NDP52^GE^ binds LC3B (Figure S1B) and LC3C (Figure 1A) more efficiently than NDP52^WT^ with statistical significance. As the interaction with LC3C was the strongest among the ATG8 family members, we focused on that. We confirmed the ability of NDP52^GE^ to bind LC3C more efficiently than NDP52^WT^ by performing co-immunoprecipitation in the opposite direction (Figure 1B), Moreover, we evaluated the colocalization between LC3C and NDP52^WT^ or NDP52^GE^ by immunofluorescence coupled with confocal microscopy analysis. The result, shown in Figure 1C, indicated a stronger colocalization between LC3C and NDP52^GE^ compared to NDP52^WT^. To reinforce these data, we moved to a semi-endogenous level by taking advantage of a HeLa cell line endogenously expressing HA-tagged LC3C [17]. By performing *in situ* Proximity Ligation Assay (PLA) we showed that the number of LC3C-NDP52^GE^ interactions was significantly greater than the number of LC3C-NDP52^WT^ interactions (Fig 1D). Finally, we assessed whether the enhanced interaction between NDP52^GE^ and LC3C required the cLIR motif alone or the contribution of other NDP52 motifs. In particular, for each NDP52 protein we generated two mutants in which either the cLIR motif (V136S) or the LIR-like motif (203-AAAA-206, ι1LIR) was functionally inactivated. These are indeed the motifs of NDP52 respectively involved in the binding to LC3C and the other ATG8 family members [14,16] and interestingly are both close to G140E substitution (Figure 2A). We performed Co-IP experiments to test the ability of these mutants to bind LC3C. The results showed that both NDP52^WT^ and NDP52^GE^ proteins mutated in the cLIR motif failed to co-precipitate LC3C (figure 2B). On the contrary, NDP52 proteins mutated in the LIR-like motif maintained the ability to co-precipitate LC3C and, similarly to NDP52^GE^, the NDP52^GE_ι1LIR^ mutant binds LC3C more efficiently than NDP52^WT_ι1LIR^ mutant (Figure 2C).

**Figure 1.**
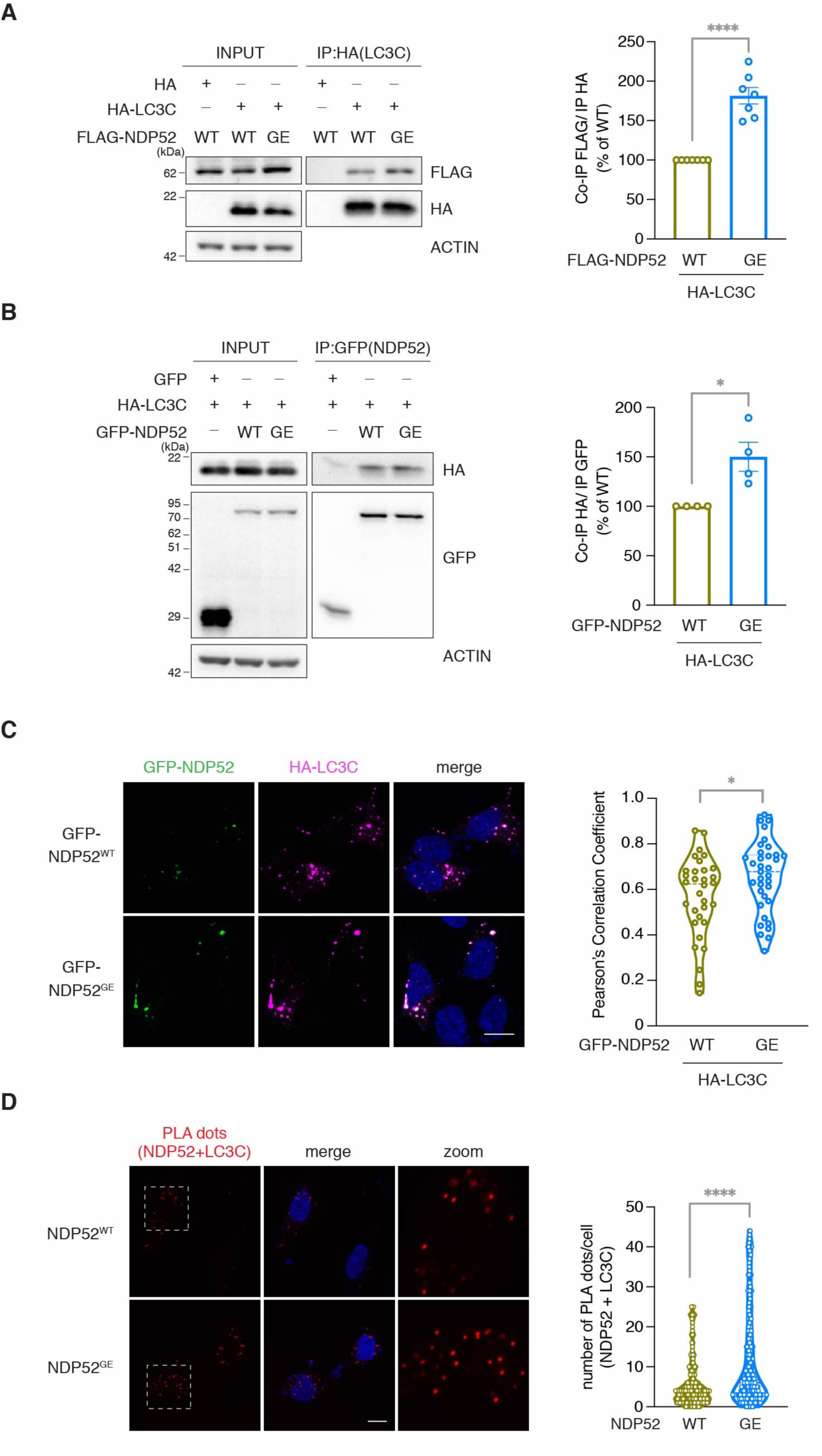
NDP52^GE^ binds LC3C more efficiently than NDP52^WT^ in a human neuroblastoma cell line. (**A**) Lysates of SH-SY5Y cells expressing the indicated FLAG- and HA-tagged proteins were immunoprecipitated with anti-HA beads. Samples were analyzed by Western blot using the indicated antibodies. The graph reports the amount of the indicated FLAG-NDP52 protein coprecipitated by the corresponding HA-LC3C protein. Data were expressed as percentage variation over FLAG-NDP52^WT^. Images and data are representative of seven independent experiments. (**B**) Lysates of SH-SY5Y cells expressing the indicated GFP- and HA-tagged proteins were immunoprecipitated with anti-GFP beads. Samples were analyzed by Western blot using the indicated antibodies. The graph reports the amount of HA-LC3C coprecipitated by the corresponding GFP-NDP52 protein. Data were expressed as percentage variation over GFP-NDP52^WT^. Images and data are representative of four independent experiments. (**C**) Representative confocal images of SH-SY5Y cells expressing the indicated GFP- and HA-tagged proteins, fixed and stained with anti-HA (magenta staining) antibodies and with DAPI (blue staining in the merge panels) to detect nuclei. Colocalization of GFP-NDP52 and HA-LC3C was quantified by measuring the Pearson’s Correlation Coefficient. The results are reported in the graph. Images are representative of three independent experiments and at least 30 cells for each condition were analyzed. Scale bar: 10μm. In B and C data are presented as means ± SEM. *p<0,05; ***p = <0,001 (two tailed unpaired *t*-test). (**D**) Representative confocal images of LC3C/NDP52 *in situ* PLA signal. HeLa cell endogenously expressing HA-LC3C were transfected with GFP-NDP52^WT^ or GFP-NDP52^GE^. *in situ* PLA experiment was performed using mouse anti-HA + rabbit anti-NDP52 primary antibodies. Dots were counted using CellProfiler software and results were shown in the violin plot graph. At least 100 cells for each condition were analyzed. ****p= <0,0001 (two tailed unpaired *t*-test). Scale bar: 10μm.

**Figure 2.**
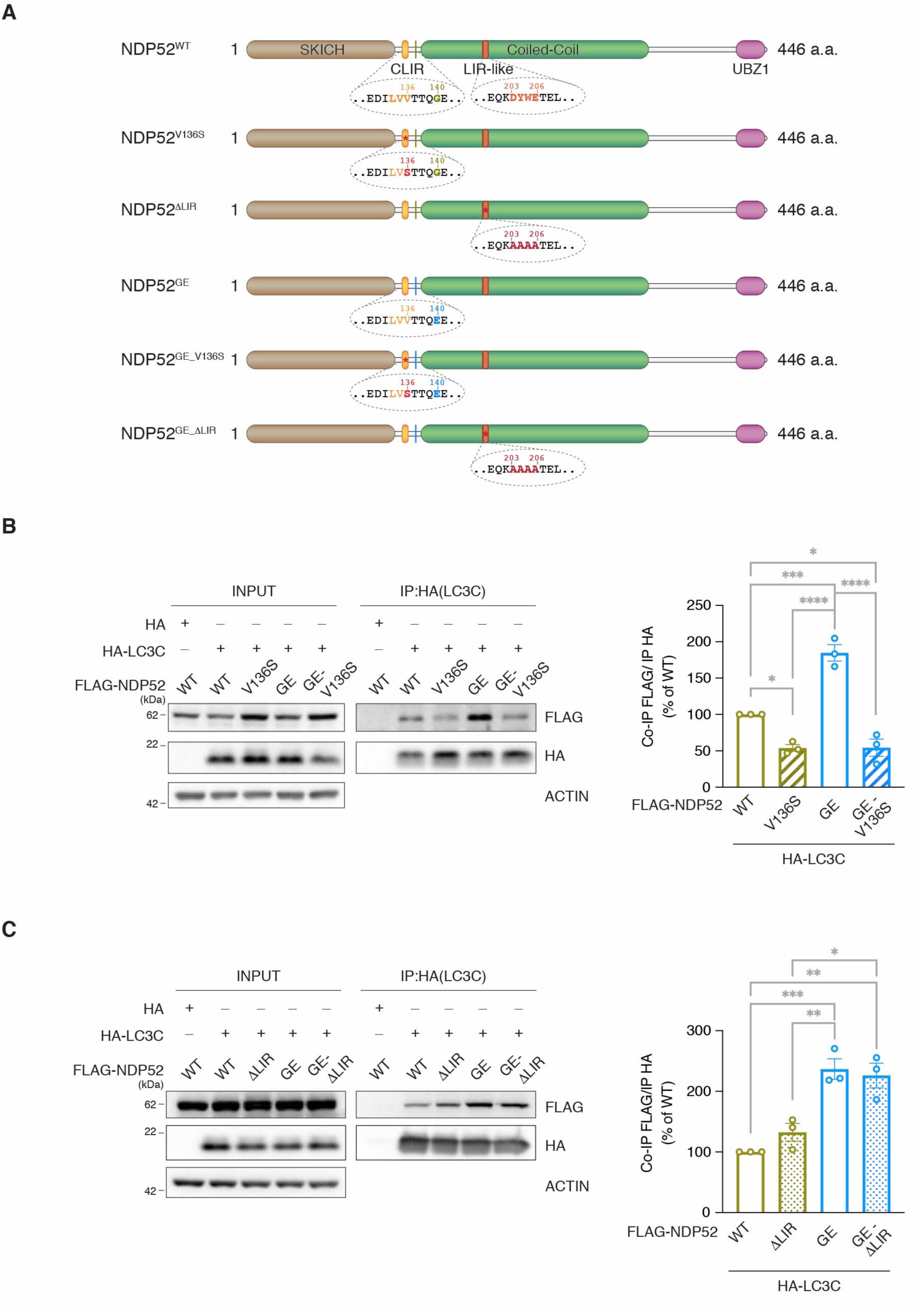
The increased binding affinity of NDP52^GE^ towards LC3C strictly depends on the cLIR motif. (**A**) Schematic representation of the human NDP52^WT^ protein, the human variant NDP52^GE^ and the NDP52 mutants used in this study. The G140E aminoacidic substitution is highlighted, and the position relative to cLIR motif as well as the LIR-like motif are shown in the expanded dotted area. SKICH = Skeletal muscle and Kidney-enriched Inositol phosphatase Carboxyl Homology; cLIR= non-canonical LC3-interacting region; LIR= LC3-interacting region; UBZ= Ubiquitin-Binding Zinc finger. (**B**) and (**C**) Lysates of SH-SY5Y cells expressing the indicated FLAG- and HA-tagged proteins were immunoprecipitated with anti-HA (B) or anti-FLAG (C) beads. Samples were analyzed by Western blot using the indicated antibodies. The graphs report the amount of the indicated FLAG-NDP52 protein coprecipitated by the corresponding HA-LC3C protein. Data were expressed as percentage variation over FLAG-NDP52^WT^. Images and data are representative of three independent experiments. ****pvalue<0,0001; *** pvalue <0,001; **pvalue < 0,01: * pvalue <0,05 (Ordinary One-way ANOVA, Turkey’s multiple comparisons test).

These data indicate that also in neuronal-like cells the NDP52^GE^ variant interacts with LC3C more efficiently than NDP52^WT^ and that this increased binding affinity is strictly dependent on the cLIR motif of NDP52.

### NDP52^GE^ delays, better than NDP52^WT^, the accumulation of hyper-phosphorylated forms of TAU in OkA treated neuronal-like cells

Since we confirmed that NDP52^GE^ binds LC3C more efficiently than NDP52^WT^, we expected that this variant is able to better recruit autophagosome, resulting in a faster degradation of cargo. Considering the previous demonstration that NDP52^WT^ facilitates the clearance of pathological hyper-phosphorylated TAU (pTAU) by autophagy [12], we wondered whether NDP52^GE^ would promote a faster degradation and in turn a slower accumulation of pTAU. In order to verify this hypothesis, we employed again the neuroblastoma SH-SY5Y cell line since it expresses all six human TAU isoforms and shows a TAU phosphorylation state similar to that of the human brain [18]. Moreover, we used Okadaic Acid (OkA), a potent pharmacological inhibitor of multiple protein phosphatases, to induce TAU hyperphosphorylation and simulate the early events of TAU-specific pathology [19–22]. We measured the levels of endogenous TAU and that of specific phosphorylated forms, reported to be highly enriched in AD brains such as pS396, pS202/pT205 (AT8) and pT212/pS214 (AT100) [23]. As expected, in basal condition (Fig. S2A, lane 0), anti-TAU signal results in several bands with different electrophoretic mobility that grouped in two regions, one around 50-60kDa and the other around 70kDa. This distribution is consistent with the molecular weights of the six TAU isoforms that range from 48kDa to 67kDa. Positive signals can also be observed for pSer396-TAU antibody (Fig. S2A, lane 0). More defined signals can be appreciated around 50-60kDa and 130-170kDa, consistent with monomeric and high molecular weight oligomeric phospho-TAU, respectively. Treatment of SH-SY5Y with 100nM OkA for 1h and 2h induced a progressive increase in the intensity and a shift in the electrophoretic mobility of the signals positive for TAU and pSer396TAU antibodies (Figure S2A). Treatment of lysates with lambda phosphatase (λPP) enzyme, that removes phosphate groups from proteins, reverted the OkA induced shift in the electrophoretic mobility of the TAU-positive signals and let disappear the bands positive for pSer396TAU antibody (Fig. S2B). Altogether these results suggest that in SH-SY5Y OkA treatment induces the formation of hyperphosphorylated TAU species, gradually and with statistical significance (Fig. S2C). The phospho-antibodies AT8 and AT100 specifically recognize disease-associated pTAU species [24–26], and, coherently, no signals can be detected in basal condition (Fig. S2D-E, lane 0). Conversely, after 1h of OkA treatment, AT8- and AT100-positive signals appeared, showing a molecular weight higher than monomeric phospho-TAU, indicative of possible TAU aggregation. The intensity of these signals increased, with statistical significance, after 2h of OkA treatment (Fig. S2D-E), proving the progressive formation of pathological oligomeric TAU species. In this context, in order to evaluate whether endogenous NDP52 directly interacts with these hyperphosphorylated forms of TAU, we performed *in situ* Proximity Ligation Assays (PLA). In particular, we tested the interaction with TAU, pSer396TAU and AT100, during OkA treatment. The results showed positive PLA dots, indicative of protein-protein interaction, for all the tested conditions (Figure S2F). Interestingly, the number of PLA dots was higher for NDP52-pSer396TAU and NDP52-AT100 couples compare to NDP52-TAU, suggesting that endogenous NDP52 might interacts more with hyperphosphorylated pSer396TAU and AT100 and less with hyperphosphorylated TAU, supporting a possible binding preference of NDP52 for certain pathological TAU species.

Afterwards, we tested whether the overexpression of NDP52^WT^ or the variant NDP52^GE^ could affect the accumulation of TAU and p-TAU species induced by OkA treatment. As shown in Figure 3, TAU accumulated more slowly in cells overexpressing NDP52 compare to control, reaching a statistically significant increase only after 2h of OkA treatment (Fig. 3A-B). A similar trend can be observed for the accumulation of pSer396-TAU (Fig. 3A-C). Interestingly, TAU accumulation is even slower in cells overexpressing NDP52^GE^ compared to NDP52^WT^, and the accumulation of pSer396-TAU did not reach a statistically significant increase even after 2h of OkA treatment (Fig. 3B-C). Thus, NDP52^GE^ appears to be more powerful than NDP52^WT^ in slowing down pTAU accumulation. No differences in the accumulation of AT8-positive pTAU can be observed in cells overexpressing NDP52^WT^ or NDP52^GE^ compared to control (Fig. 3D). On the contrary, the accumulation of AT100-positive pTAU is statistically significant lower in cell overexpressing NDP52^GE^ compared to control or to NDP52^WT^ (Fig. 3E). It should be noted that no difference can be observed between control and NDP52^WT^ overexpressing cell as concern the accumulation of AT100-positive signals. Altogether these results suggest that in this *in vitro* system, in which OkA treatment mimics the initial steps of TAU aggregation, NDP52^WT^ is able to slow down the accumulation of monomeric and high molecular weight oligomeric TAU species, positive to TAU and pSer396TAU antibodies. The variant NDP52^GE^ proves to be stronger than NDP52^WT^ in delaying TAU and pSer396TAU accumulation. Moreover, unlike NDP52^WT^, NDP52^GE^ also proved effective in slowing the accumulation of AT100-positive pTAU, which is one of the most specific correlates of toxicity in AD [27].

**Figure 3.**
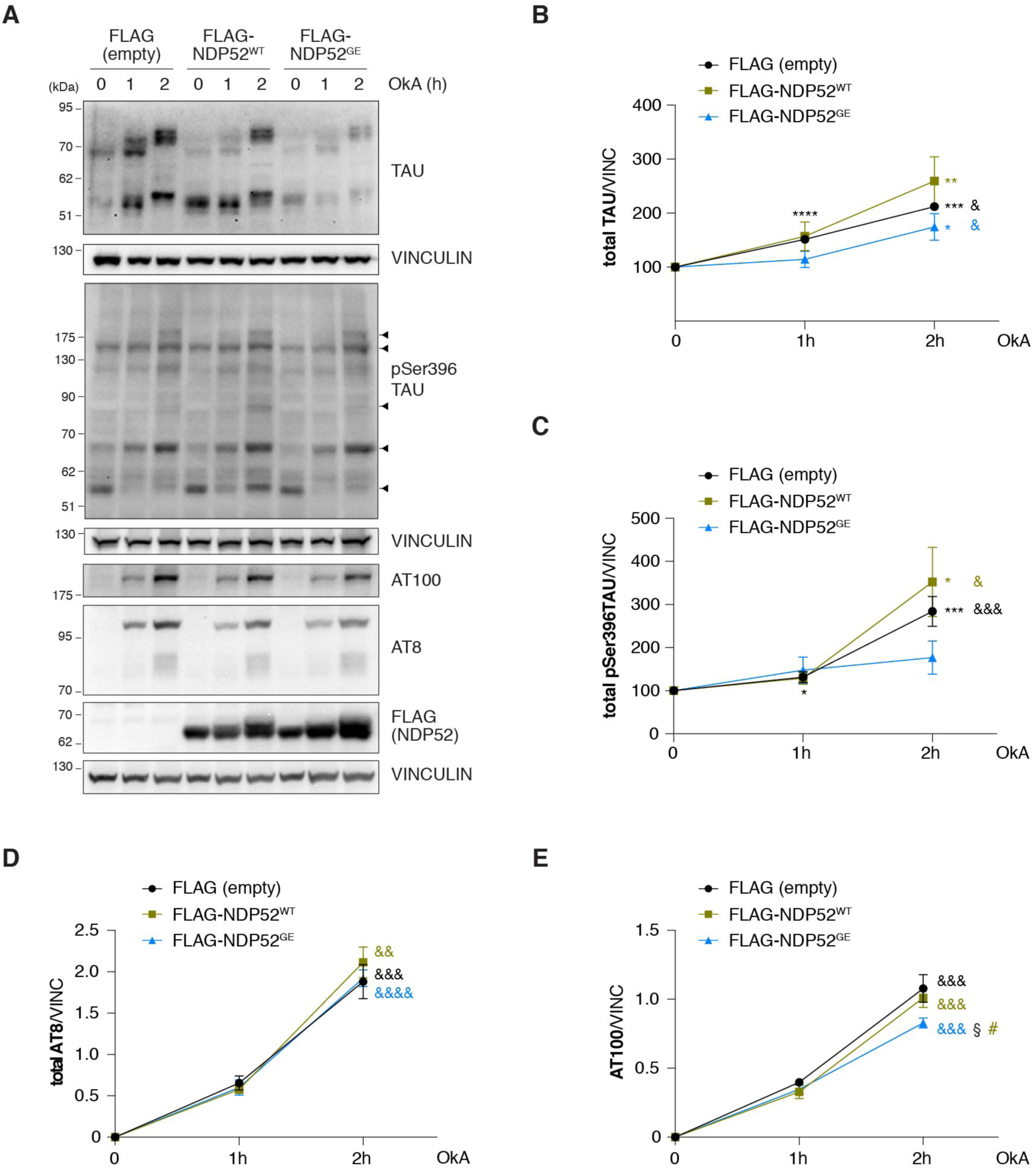
NDP52^GE^ reduces pTAU levels better than NDP52^WT^, in a human neuroblastoma cell line. (**A**) SH-SY5Y cells expressing the indicated FLAG-tagged proteins were treated with 100nM OkA for the indicated times. Western blot analysis was performed using the indicated antibodies. Images are representative of eight independent experiments. Arrow heads indicate pSer396TAU specific bands used for densitometry analysis. (**B and C**). Signals from TAU antibodies were measured and normalized to the corresponding signal of VINCULIN, used as loading control. Results are expressed as percentage variation of the levels measured at time 0 for each FLAG-tagged protein expressing group. Data are representative of eight independent experiments. (**D and E**). Signals from TAU antibodies were measured and normalized to the corresponding signal of VINCULIN, used as loading control. Data are representative of eight independent experiments. Data information: In (B-E) data are reported as means ± SEM. * indicates comparison to time 0 of each FLAG-tagged protein expressing group: ****pvalue<0,0001; *** pvalue <0,001; * pvalue <0,05. & indicates comparison between 1h and 2h of each group: &&&& pvalue <0,0001; &&& pvalue <0,001, & pvalue <0,05. § indicates comparison between FLAG-NDP52^GE^ 2h and empty 2h: § pvalue <0,05. # indicates comparison between FLAG-NDP52^GE^ 2h and FLAG-NDP52^WT^ 2h: # pvalue <0,05 (two tailed unpaired *t*-test).

### NDP52^WT^ and NDP52^GE^ bind hyperphosphorylated TAU, pSer396TAU and pathological AT100-positive TAU with similar efficiency

We have shown that NDP52^GE^ binds the autophagic machinery (LC3C but also LC3B) more efficiently than NDP52^WT^, which supports its greater ability in eliminating TAU. However, at this stage, we could not exclude the possibility that NDP52^GE^ was also able to bind TAU more efficiently than NDP52^WT^. To address this point, we examined the interaction of NDP52 proteins with TAU or the hyperphosphorylated forms of TAU. We performed *in situ* PLA experiments on SH-SY5Y cells overexpressing NDP52^WT^ or NDP52^GE^ in basal condition or upon OkA treatment. In basal condition, the results showed PLA dots for both NDP52^WT^ and NDP52^GE^, suggesting that interactions with TAU also occur at a non-pathological level. The number of PLA dots did not significantly differ between cells overexpressing NDP52^GE^ or NDP52^WT^, suggesting that, in basal condition, NDP52 proteins bind TAU with similar efficiency. Upon OkA treatment, PLA dots can be observed for all the couples tested (Fig 4) and again the number of PLA dots was not significantly different in cells overexpressing NDP52^GE^ or NDP52^WT^. This suggests that the hyperphosphorylated forms of TAU were bound by both NDP52 proteins with similar efficiency. Compared to the basal condition, the number of PLA dots increased upon OkA treatment. However, this increase was statistically significant in cells overexpressing NDP52^WT^ but not in those overexpressing NDP52^GE^. This could be explained by the fact that NDP52^GE^ degrades the hyperphosphorylated forms of TAU more efficiently than NDP52^WT^ (Fig 3) so that the NDP52^GE^/phospho-TAU complexes were likely removed faster than those with NDP52^WT^. Even though further experiments will be required to better investigate this latter point, our results clearly indicate that NDP52^WT^ and NDP52^GE^ do not differ in their binding affinity to TAU or its hyperphosphorylated forms.

**Figure 4.**
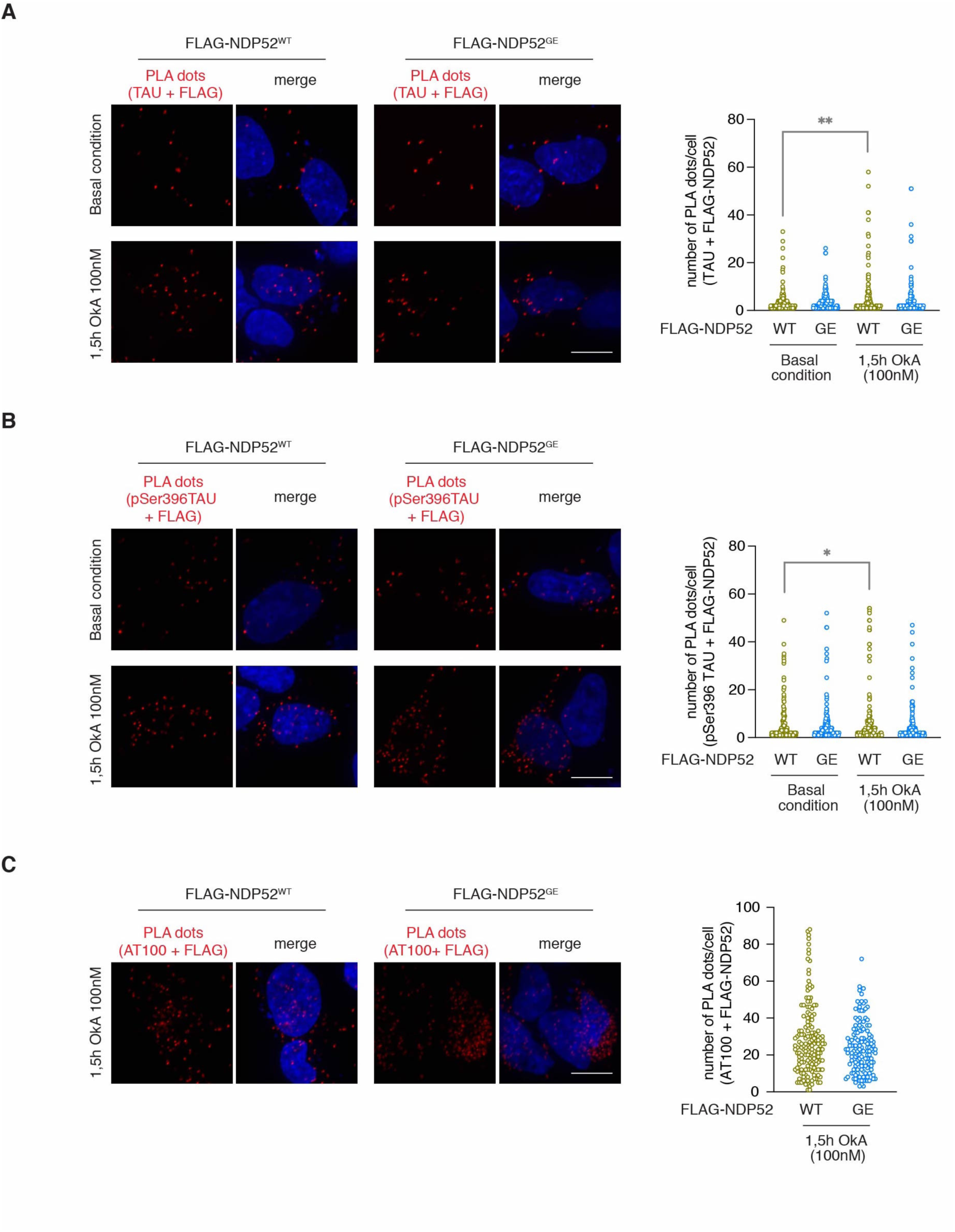
NDP52^WT^ and NDP52^GE^ bind non-pathological and pathological TAU with comparable efficiency. Representative confocal images of *in situ* PLA performed in SH-SY5Y overexpressing FLAG-NDP52^WT^ or FLAG-NDP52^GE^ and treated with DMSO (basal condition) or with 100nM OkA for 1,5 h at 37 °C. *In situ* PLA experiments was performed using anti-FLAG/anti-TAU (**A**), anti-pSer396TAU/anti-FLAG (**B**), anti-FLAG/anti-AT100 (**C**) primary antibodies. Dots were counted using CellProfiler software and results were shown in the violin plot graphs. At least 100 cells for each condition were analyzed. In A and B: **pvalue < 0,01; * pvalue <0,05 (Ordinary One-way ANOVA, Turkey’s multiple comparisons test). In C: not statistical (two tailed unpaired *t*-test). Scale bar: 10μm.

### PP2A dephosphorylates NDP52 *in vitro*

In the context of selective autophagy, phosphorylation(s) of cargo receptors is required to promote their interaction with ATG8 family proteins [28,29]. On the other hand, phosphatases control these phosphorylation sites in order to modulate the autophagic process [30]. We thus wondered whether NDP52 may be regulated by such modification during the removal of TAU. To this end, we analyzed the electrophoretic mobility of endogenous NDP52 from extracts of SH-SY5Y cells treated with OkA. As shown in Figure 5A, upon 1h of OkA treatment, we observed the appearance of a slower migrating band recognized by the NDP52 antibody. Moreover, the intensity of this band increased upon 2h of OkA treatment. To verify if this signal corresponds to a phosphorylated form of endogenous NDP52, cell lysates were treated with the λ phosphatase (λPP). As shown in Figure 5B, the slower migrating band of NDP52 specifically disappeared in the λPP-treated samples, strongly suggesting that it corresponds to a phosphorylated form of endogenous NDP52. In general, phosphorylation is regulated by the interplay between kinases and phosphatases. Thus, our result that OkA treatment induces the accumulation of phospho-NDP52 has two important implications. On one hand, it confirms that, as other autophagic receptor, NDP52 could be phosphorylated when the protein is active in selective autophagy [28,29,31]. On the other hand, it indicates that NDP52 is dephosphorylated by, at least, one of the phosphatases inhibited by OkA. Among them, Protein Phosphatase 2A (PP2A) is the primary TAU phosphatase in the human brain [32], and plays a crucial role in the AD process [33]. Thus, we wondered whether NDP52 could be a new direct target of PP2A.

**Figure 5.**
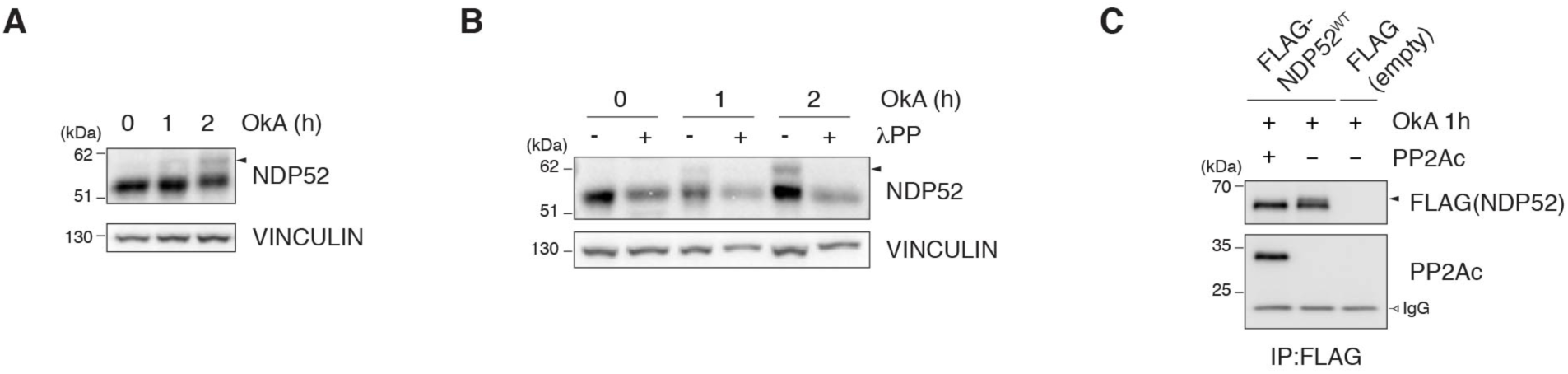
Protein Phosphatase 2A (PP2A) dephosphorylates NDP52 *in vitro*. (**A**) Lysates of SH-SY5Y cells treated with 100 nM Okadaic Acid (OkA) for the indicated time were analyzed by western blot using the indicated antibodies. Arrow head indicates the supposed phosphorylation of NDP52. Images are representative of three independent experiments. (**B**) Lysates of SH-SY5Y cells treated with 100nM OkA for the indicated time were split and incubated (+) or not (-) with Lambda Protein Phosphatase (λPP). Samples were analyzed by western blot using the indicated antibodies. Arrow head indicates the phosphorylation of NDP52 that disappeared in λPP treated samples. Images are representative of three independent experiments. (**C**) *In vitro* dephosphorylation assay. SH-SY5Y cells expressing FLAG empty or FLAG-NDP52^WT^ proteins were treated with 100nM OkA for 1h. Lysates were immunoprecipitated with anti-FLAG beads and then split and incubated with (+) or without (-) the recombinant purified catalytic subunit of PP2A (PP2Ac). Samples were then analyzed by Western blot using the indicated antibodies. Immunoprecipitated FLAG empty sample treated with OkA but not with PP2Ac (last lane) was used as negative control. IgG are shown as loading control. Images are representative of three independent experiments.

To verify this hypothesis, we performed an *in vitro* dephosphorylation assay using the purified recombinant PP2A catalytic subunit (PP2Ac). As shown in Figure 5C, the slower NDP52-migrating band, observed after OkA treatment (lane 2), disappeared in the presence of PP2Ac, indicating that PP2A directly dephosphorylates NDP52. We also tested the variant NDP52^GE^ by the *in vitro* dephosphorylation assay (Fig. S3), confirming that both NDP52^WT^ and NDP52^GE^ are direct targets of PP2A.

### NDP52^GE^ mediates a stronger autophagic removal of pathological P301S-TAU compared to NDP52^WT^

In order to further investigate the involvement of NDP52 in the clearance of pathological TAU we took advantage of a different *in vitro* system, that is a P301S mutant TAU inducible SH-SY5Y cell line [34]. Firstly, we confirmed the induction of P301S-TAU expression upon tetracycline treatment (Fig 6A and 6B) and monitored the proteins levels of endogenous LC3 and NDP52. Interestingly, both LC3II/LC3I ratio and endogenous NDP52 protein level increased upon tetracycline treatment (Fig 6B) suggesting that the expression of P301S-TAU increases autophagosome formation and activates the autophagy pathway, possibly involving NDP52.

**Figure 6.**
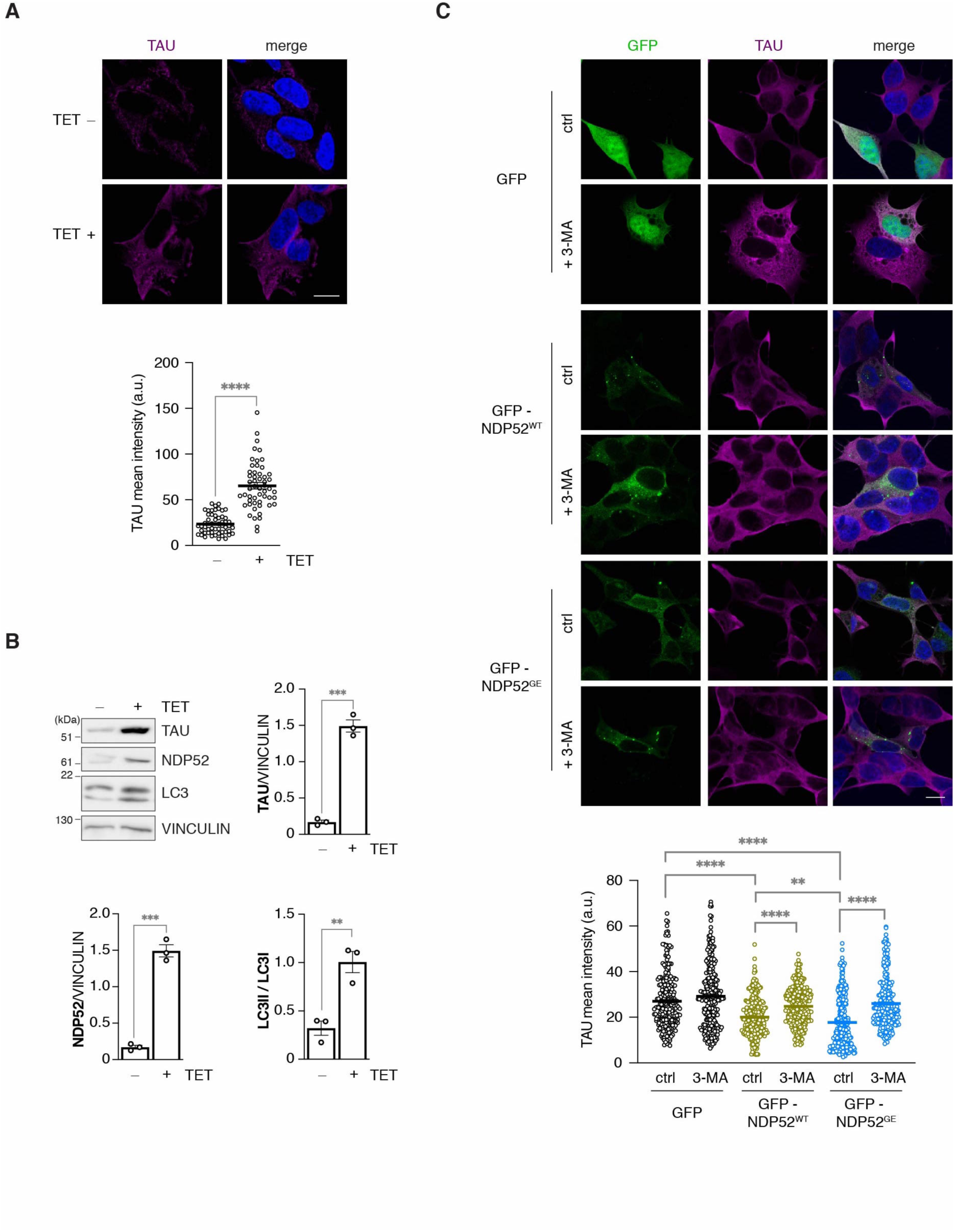
NDP52^GE^ is more efficient than NDP52^WT^ to mediate autophagic removal of P301S-TAU. (**A**) Representative confocal images of human SH-SY5Y P301S-TAU inducible cells non-treated (TET -) or treated (TET +) with tetracycline for 24h to induce P301S-TAU expression, fixed and stained with anti-TAU (magenta staining) antibody and with DAPI (blue staining in the merge panels) to detect nuclei. Intensity of anti-TAU signal was measured and results are reported in the graph. Images are representative of three independent experiments and at least 100 cells for each condition were analyzed. ****p= <0,0001 (two tailed unpaired *t*-test). Scale bar: 10μm. (**B**) Western blot analysis of cells treated as in (A). Images are representative of three independent experiments. Signals from TAU and NDP52 antibodies were measured and normalized to the corresponding signal of VINCULIN, used as loading control. Signals from lipidated (LC3II, faster migrating LC3 band) and unlipidated (LC3I, slower migrating LC3 band) LC3 were measured and LC3II/LC3I ratio was calculated. Data are reported as means ± SEM. *** pvalue <0,001; **pvalue < 0,01 (two tailed unpaired *t*-test). (**C**) Representative confocal images of human SH-SY5Y P301S-TAU inducible cells overexpressing GFP-NDP52^WT^ or GFP-NDP52^GE^ and treated with tetracycline to induce P301S-TAU expression. After 16h, medium was substituted with a TET-free medium and cells were left untreated (ctrl) or were treated with the autophagic inhibitor 3-Methyladenine (+ 3-MA) for 8h. Cells were then fixed and stained with anti-TAU (magenta staining) antibody and with DAPI (blue staining in the merge panels) to detect nuclei. Intensity of anti-TAU signal was measured and results are reported in the graph. Images are representative of three independent experiments and at least 100 cells for each condition were analyzed. **** pvalue <0,0001; **pvalue < 0,01 (two tailed unpaired *t*-test).

Our previous results showed that NDP52^WT^ and NDP52^GE^ slowed down the accumulation of early pathological pTAU. However, in the OkA-treated system we cannot distinguished whether TAU expression was reduces and/or its turnover was enhanced. To assess this point, GFP (as control) or GFP-NDP52^WT^ or GFP-NDP52^GE^ constructs were overexpressed and P301S-TAU was induced by tetracycline (TET) treatment in inducible SH-SY5Y cells. Upon 16h, medium was replaced by TET-free medium to stop P301S-TAU expression and, 8h later, the levels of TAU were measured by immunofluorescence assay coupled with confocal microscopy analysis. As shown in figure 6C NDP52^WT^ overexpression significantly reduces the level of P301S-TAU compared to control. Moreover, NDP52^GE^ variant exerts a more efficient role in mediating this process compared to NDP52^WT^. This result suggests that NDP52 exerts its role by possibly enhancing TAU degradation, rather than affecting TAU expression. To verify this hypothesis, cells were treated with 3-Mehyladenine (3-MA), a well-known autophagy inhibitor. In this case, both NDP52^WT^ and NDP52^GE^ failed to reduce the level of P301S-TAU (figure 6C), confirming that NDP52 proteins mediates TAU clearance through the autophagy process.

### Human NDP52 ameliorates TAU-mediated toxicity in an *in vivo* AD model of *Drosophila melanogaster*, and the variant NDP52^GE^ proves to be more powerful than NDP52^WT^

Our previous results showed that NDP52 reduces the accumulation of pathological TAU, with the variant NDP52^GE^ being stronger than NDP52^WT^. To assess whether these effects would ameliorate the neurodegeneration induced by the accumulation of pathological TAU, we moved to an *in vivo* system. In particular we used a *Drosophila* model of TAU toxicity that expresses the longest hTAU isoform (2N4R, hereafter refers to as hTAU) under the UAS promoter and generated two transgenic fly lines expressing either human NDP52^WT^ or NDP52^GE^ cDNA, downstream of the UAS promoter. These constructs were inserted in the same genomic site to avoid any variability of expression due to the genomic environment, and we used the UAS-GAL4 system [35] to specifically express the human transgenes in different fly tissues. Firstly, we evaluated whether the expression of hNDP52^WT^ or hNDP52^GE^ modifies the ratio between the lipidated and unlipidated forms of dAtg8 (dAtg8 II/dAtg8 I ratio). Interestingly, we discovered that the expression of hNDP52^GE^ in fly eyes (under the control of GMR-GAL4 driver), increases the dAtg8 II/dAtg8 I ratio, compared to the control and to NDP52^WT^ (figure 7A). This result suggests that NDP52 could interact with Drosophila autophagic machinery, and that NDP52^GE^ is able to favor the autophagosome formation better than NDP52^WT^ also in *Drosophila Melanogaster*. Then we evaluated the effect of co-expression of hTAU and hNDP52^WT^ or NDP52^GE^. Expressing hTAU early during eye development under the control of *eyeless*-GAL4 driver resulted in retina degeneration linked to a significant reduction in the eye area (Fig. 7B, Ey> vs Ey>hTAU) [36]. This reduction is partially rescued by either NDP52^WT^ or NDP52^GE^. Interestingly, the rescue is more significant in flies expressing hNDP52^GE^. In this context we also monitored by western blot analysis the levels of TAU and pTAU (pSer396TAU, AT8 and AT100) proteins. No signals were detected for AT100 and AT8 antibodies.

**Figure 7.**
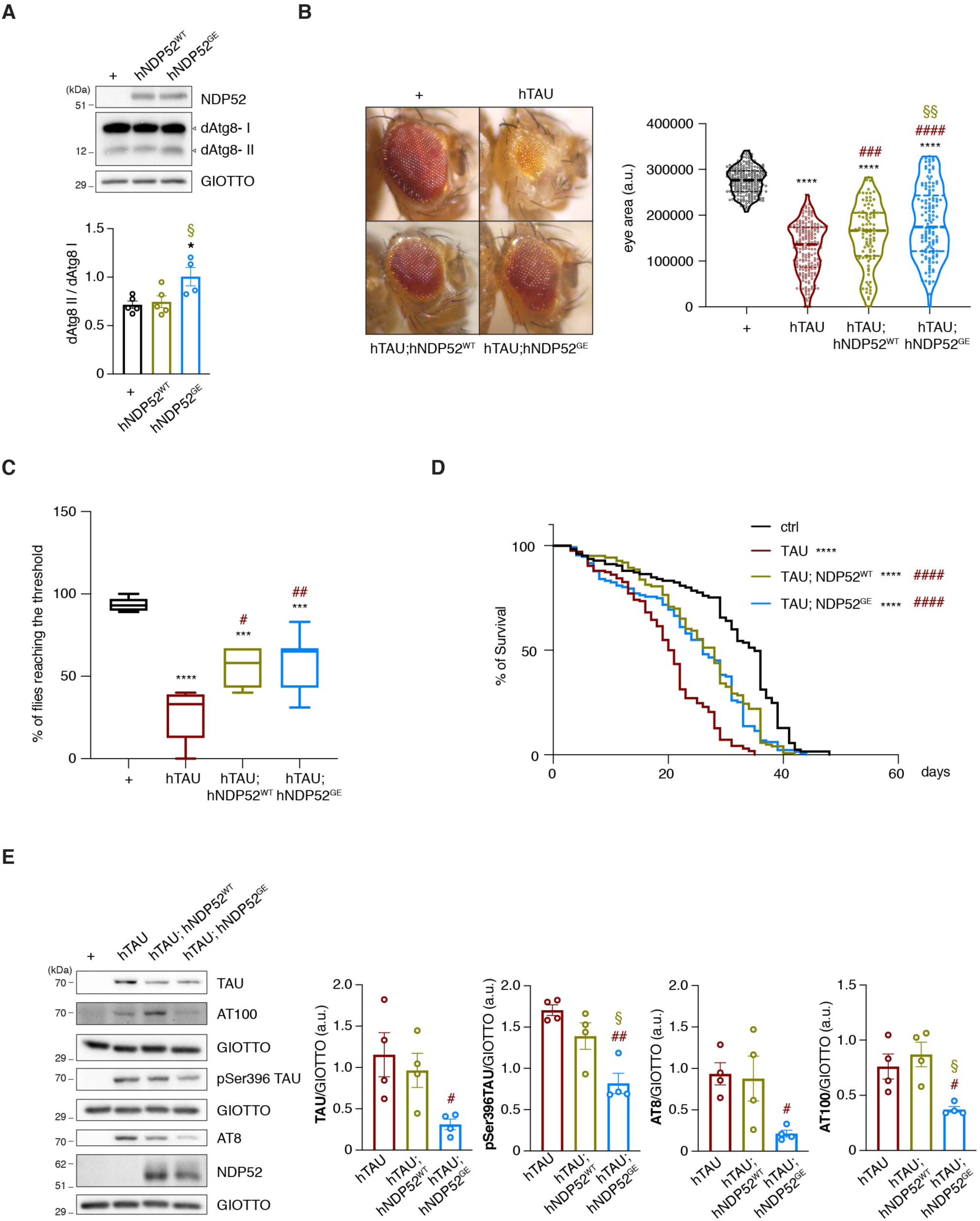
hNDP52 partially rescues hTAU phenotypes in *Drosophila melanogaster* and the variant NDP52^GE^ proves to be more powerful compared to NDP52^WT^. (A) Lysates from heads of flies expressing the indicated transgenes in the eye, at 29°C, under control of GMR-GAL4 driver (GMR>) were analyzed by western blot using the indicated antibodies. + refers to non-transgenic flies used as control. Images are representative of 4 independent experiments. The ratio between dAtg8II/dAtg8I was calculated and reported in the graph as means ± SEM. * indicates comparison of dAtg8II/dAtg8I between flies expressing hNDP52^GE^ and control: * pvalue <0,05; § indicates comparison of dAtg8II/dAtg8I between flies expressing hNDP52^GE^ and hNDP52^WT^: § pvalue <0,05 (Ordinary One-way ANOVA, Turkey’s multiple comparisons test). (**B**) Representative images of eye from flies expressing the indicated human (h) transgenes at 25°C under control of eyeless-GAL4 driver (Ey>). + indicates non-transgenic flies used as control. Violin plot below reports eye areas. Median and quartiles are shown as dotted lines. More than 150 eyes from both males and females flies were measured for each condition. * indicates comparison to control (+): **** pvalue <0,0001; # indicates comparison to hTAU: ### pvalue <0,001, #### pvalue <0,0001; § indicates comparison to hTAU;hNDP52^WT^: §§ pvalue <0,01 (Ordinary One-way ANOVA, Turkey’s multiple comparisons test). (**C**) Box and whiskers (min to max) plot showing the percentage of flies reaching the threshold. Female flies expressed panneuronally (elav-GAL4, ELAV>) the indicated human transgenes at 29°C. + indicates non-transgenic flies used as control. More than 40 flies per genotype were analyzed (Control n=54 flies; hTAU n=44; hTAU, NDP52^WT^ n=53; hTAU, NDP52^GE^ n=48 flies). * indicates comparison to control (+): **** pvalue <0,0001; *** pvalue <0,001; # indicates comparison to hTAU: # pvalue <0,05, ## pvalue < 0,01 (Ordinary One-way ANOVA, Turkey’s multiple comparisons test). (**D**) Survival curve showing lifespan of female flies expressing panneuronally (elav-GAL4, ELAV>) the indicated transgenes at 29°C. * indicates comparison to control (+): **** pvalue <0,0001; # indicates comparison to hTAU: #### pvalue <0,0001 (Log-rank (Mantel-Cox) test). (**E**) Lysates from heads of flies expressing panneuronally (elav-GAL4, ELAV>) the indicated transgenes at 29°C were analyzed by western blot using the indicated antibodies. + refers to non-transgenic flies used as control. Images are representative of 4 independent experiments. Signals from the indicated TAU antibodies were measured and normalized to the corresponding signal of GIOTTO, used as loading control. Results, expressed as means ± SEM, are shown in the graphs below. # indicates comparison to hTAU: # pvalue < 0,05, ## pvalue < 0,005; § indicates comparison to hTAU;hNDP52^WT^: § pvalue < 0,05 (Ordinary One-way ANOVA, Turkey’s multiple comparisons test).

Interestingly, the levels of pSer396TAU were reduced, with statistical significance, in the strains co-expressing hTAU with both human variants of NDP52 compared to the strain only expressing hTAU. Moreover, the level of hTAU protein was lower in the strain co-expressing hTAU and hNDP52^GE^, compared to the strain only expressing hTAU (Fig S4A). This strongly suggests that NDP52^GE^ counteracts TAU accumulation more efficiently than NDP52^WT^.

To further explore the beneficial effect of hNDP52 expression on hTAU-induced toxicity, we performed a locomotor assay using a panneuronal driver (*elav*-GAL4). Flies expressing hTAU showed a statistically significant reduction of locomotor ability compared to control (Figure 7C). Flies expressing either NDP52^WT^ or NDP52^GE^, showed a partial rescue of the climbing activity. Even though not statistically significant, a slight improvement in the climbing performance was observed in flies expressing hNDP52^GE^ compared to hNDP52^WT^. In addition, as the neurodegeneration strongly affects fly viability, we performed a lifespan assay to compare hTAU toxicity in the different *Drosophila* strains. The results showed that the lifespan of both females (fig 7D) and males (figS4B) flies co-expressing hNDP52^WT^ or hNDP52^GE^ significantly increases compared with flies expressing only hTAU.

Finally, we tested whether the co-expression of hNDP52^WT^ or hNDP52^GE^ affected the protein levels of TAU and pTAU species in fly neurons. Although the expression of NDP52^WT^ was not sufficient to produce a statistically significant reduction of TAU and pTAU protein levels (Fig. 7E), the expression of NDP52^GE^ showed a statistically significant reduction in the levels of TAU and pTAU (pSer396TAU, AT8 and AT100) compared to controls. Moreover, flies expressing NDP52^GE^ showed a statistically significant reduction of pSer396TAU and AT100 compared to flies expressing NDP52^WT^.

Collectively, our data showed for the first time, that NDP52 mitigates hTAU-induced neurotoxicity *in vivo*. Moreover, they showed that NDP52^GE^ promotes a better rescue of the eye phenotype and a significant reduction of TAU and pathological pTAU protein levels. In addition, considering that the way in which NDP52 transgenic fly lines were generated ensures the same level of expression and that in all the tested conditions NDP52^GE^ protein levels were comparable to those of NDP52^WT^ (figS4A-C-D), our data strongly suggest that the more beneficial effect of NDP52^GE^ does not result from an increase in the protein’s stability but rather from its greater capacity to promote the autophagy-mediated elimination of TAU proteins.

### Worldwide frequency of the variant allele coding for NDP52^GE^ positively correlates with the incidence and the prevalence of AD and other dementia

Our results strongly support the variant NDP52^GE^ as a protective factor for AD. By analyzing available data reported on 1000Genomes [37], we found that the variant allele coding for NDP52^GE^ (c.491G>A, rs550510, p.G140E, hereafter named allele A) has a global frequency of 24,3%. However, its frequency shows a strikingly variable distribution among different populations. In particular, allele A has the minimum frequency in the African continent (2,5%), being completely absent in West Africa (Nigeria). On the contrary, it is very common in the East Asian populations, where it reaches a frequency of 65,4% (Figure 8A). Interestingly, according to the worldwide epidemiological investigation of AD, these regions - Africa and East Asia - are also the ones that show respectively the lower and the higher rates of prevalence of AD [38] (Figure 8A). The widespread global distribution of this variable aligns with the observation that common genetic variants often have pleiotropic effects, which vary based on interactions with factors related to specific populations, including other genetic characteristics and environmental conditions. Thus, the higher prevalence of allele A in East Asia might represent a consequence of an advantage provided by NDP52^GE^. In order to investigate this hypothesis, we analyzed the correlation between the worldwide frequency of the allele A and the incidence or prevalence of AD and other dementia [39]. On the basis of 2019 Global Burden Disease (GBD) database, Li et al. calculated the trends of age standardized Incidence or Prevalence rates, from 1999 to 2019, in 21 GBD regions. Among these 21 GBD regions, divided on the basis of geographic location, we selected those for which population the 1000 Genomes database reports genetic data and then performed correlation analysis. The results of the statistical analysis, shown in Figure 8B, indicate that the frequency of the allele A was significantly positively correlated with the incidence (r=0,8181 p=0,0038) and the prevalence (r=0,8676 p=0,0011) of AD and other dementia. To further examine this point, we performed similar analyses testing the correlation of the worldwide incidence or prevalence data with the frequency of the AA genotype, as well as the frequency of the wild type allele (allele G), the GG genotype or the heterozygous AG genotype. The frequency of the AA genotype was significantly positively correlated with the incidence (r=0,8591 p=0,0014) and the prevalence (r=0,8869 p=0,0006) of AD and other dementia (Figure 8C). Conversely, the frequency of both the wild type allele (allele G) or the GG genotype were significantly inversely correlated with the incidence (allele G: r=-0,8181 p=0,0038; genotype GG: r=-0,7742 p=0,0086) or the prevalence (allele G: r=-0,8676 p=0,0011; genotype GG: r=-0,835 p=0,0026) of AD and other dementia. No correlation was observed between the frequency of the heterozygous AG genotype and the incidence of AD (r=0,5518 p=0,0982), and just a slight positive correlation was observed between the frequency of the heterozygous AG genotype and the prevalence of AD and other dementia (r=0,6411 p=0,0458) (Figure S5). Although further analysis will be required to understand whether the allele A underwent adaptive evolution, these results may support the role of the NDP52^GE^ variant as a resilience factor in AD.

**Figure 8.**
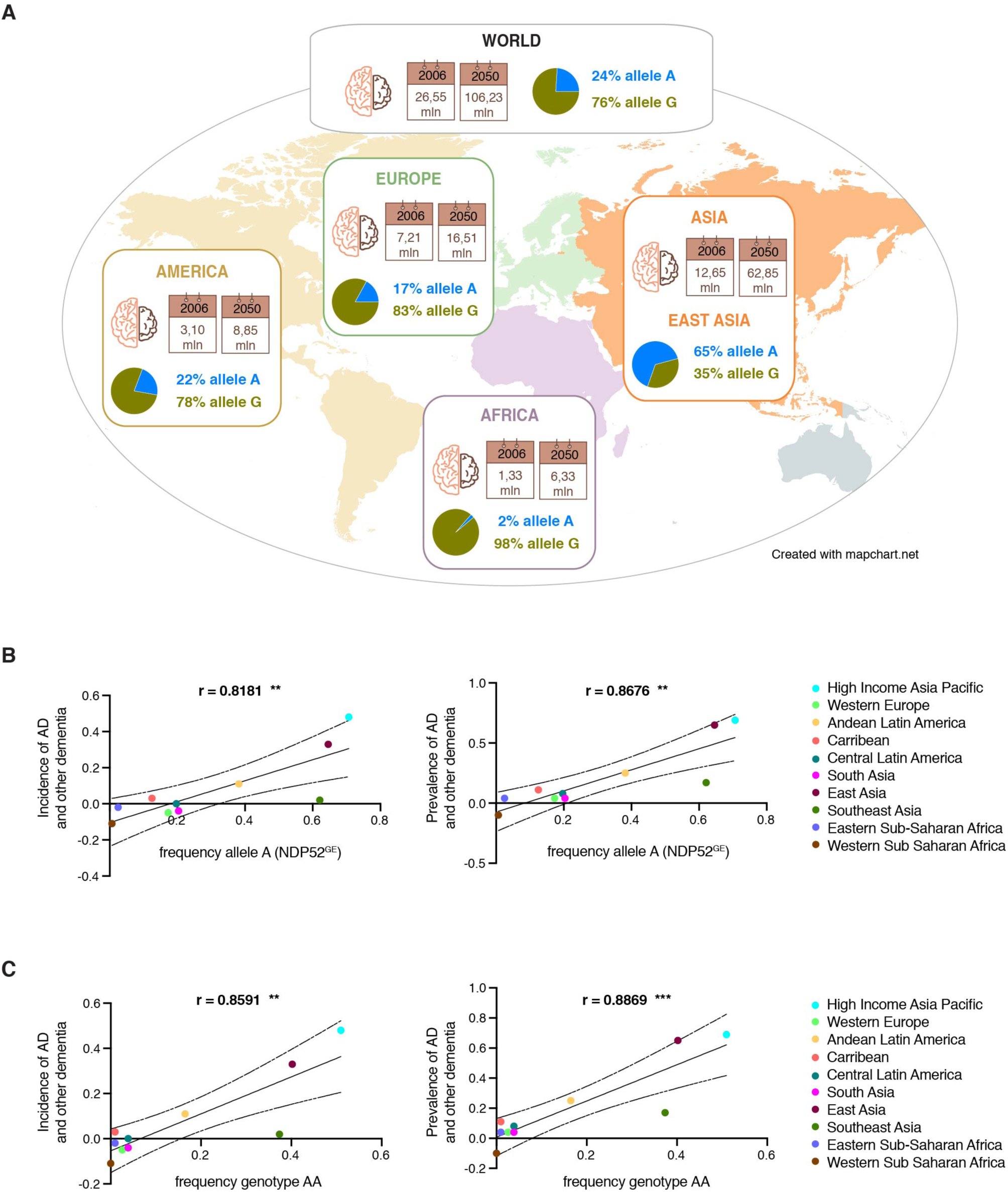
Worldwide frequency of the variant allele coding for NDP52^GE^ correlates with the incidence and the prevalence of AD and other dementia. (**A**) Cartoon showing the continental distribution of the prevalence of AD measured in 2006 and the projection for 2050 reported in [38] and the frequencies of allele A (rs550510, coding for NDP52^GE^) and allele G (coding for NDP52^WT^) reported in 1000 Genomes Project Consortium [37]. (**B and C**) Graphs showing the correlation analysis between worldwide incidence or prevalence of AD and other dementia and the frequency of allele A (B) or genotype AA (C). Incidence and prevalence data are based on the estimated annual percentage changes of age-standardized rates of incidence and prevalence from 1990 to 2019 reported in [39]. Data of allele or genotype frequencies are from the 1000 Genomes Project Consortium [37]. Pearson’s Correlation Coefficient (r) and 95% confidence interval are shown. * pvalue (two-tailed) < 0,05; ** pvalue (two-tailed) <0,005. Data information: For (B-C) correlation analyses were performed with Prism8 software.

## DISCUSSION

In this study, we characterized the contribution of NDP52^GE^, a variant of the autophagic receptor NDP52/CALCOCO2 (c.491G>A, rs550510, p.G140E), to mitigate AD by efficiently clearing, through autophagy, pathological and hyperphosphorylated TAU proteins (Figure 9).

**Figure 9.**
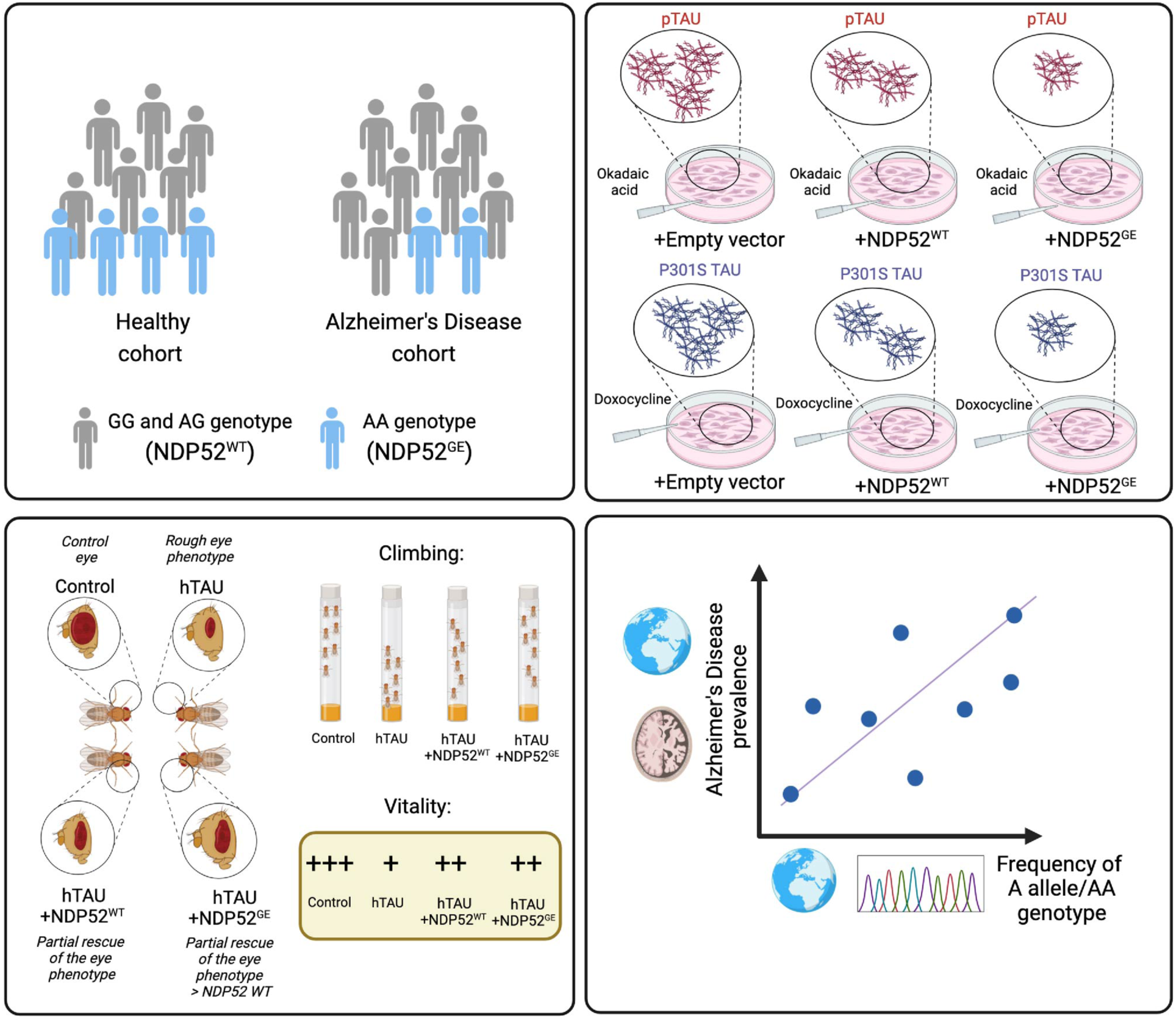
NDP52^GE^ is a protective factor in AD and counteracts pTAU accumulation. Illustration of the highlights of the work. Using a genetic analysis, we showed that NDP52^GE^ is a protective factor in AD. We demonstrated that this variant counteracts the accumulation of pathological pTAU better than NDP52^WT^, both *in vitro* and *in vivo*. Furthermore, we showed in a *Drosophila melanogaster* model of TAU-mediated AD that NDP52^GE^ partially rescued different pathological phenotypes. Finally, we found an interesting positive correlation between the worldwide distribution of the allele coding NDP52^GE^ and the incidence and prevalence of AD and other dementia. Created with BioRender.com.

Autophagy appears to be the primary route of clearance of TAU in healthy neurons and its dysfunction leads to the accumulation of TAU oligomers and insoluble aggregates [40]. Growing evidence suggests that these species are the most toxic for neurons [9–11] and strongly correlate with the synaptic dysfunction underlying memory deficit in AD [41,42]. On the contrary, induction of autophagy can significantly reduce the formation of these species and thus could offer protection against TAU pathogenicity [40,43,44]. In addition, accumulating evidence shows that autophagy is required for synaptic functions, including neurotransmission and synaptic plasticity. [45,46]. Preserved autophagy, accompanied with significantly reduced TAU pathology and maintenance of cognitive integrity have been observed in resilient individuals who have AD neuropathology but remain non-demented [47]. In these individuals, autophagy may be responsible for the maintenance of cognitive integrity through the efficient removal of TAU oligomers, supporting the potential of autophagy-inducing strategies in AD therapeutics. Interestingly, in the hippocampus of these resilient AD patients, the protein levels of the autophagic receptor NDP52/CALCOCO2 results increased and negatively correlates to TAU oligomers [47]. Moreover, NDP52 has been shown to target pathological hyper-phosphorylated TAU to the autophagic degradative pathway, so that it might contribute to the prevention of TAU aggregation by acting as an autophagy receptor that promotes TAU clearance [12].

Consistent with literature, in this study we demonstrated that NDP52 directly binds hyperphosphorylated pathological forms of TAU and promotes their degradation through autophagy. More interestingly, we showed that NDP52^GE^, a variant of NDP52, is significantly associated with a decreased susceptibility to AD, resulting a protective factor for it.

We hypothesized that NDP52^GE^ mitigates AD by clearing hyperphosphorylated and pathological TAU proteins more efficiently than NDP52^WT^. In support of this, we showed in two different *in vitro* systems that NDP52^GE^ reduced the accumulation of pathological P301S-TAU and hyperphosphorylated forms of TAU (pSer396, AT8 and AT100) better than NDP52^WT^. By taking advantage of a *Drosophila melanogaster in vivo* model of TAU-mediated AD, we demonstrated that NDP52 mitigates hTAU-induced neurotoxicity, and we confirmed a prominent effect of NDP52^GE^ compared to NDP52^WT^.

Mechanistically, we anticipated that the NDP52^GE^ variant was able to “clean” pathological hyper-phosphorylated TAU better than NDP52^WT^ through a stronger autophagy-mediated degradation. Indeed, our data indicate that NDP52^WT^ and NDP52^GE^ do not differ in the binding affinity to TAU or hyperphosphorylated TAU. On the contrary, we confirmed that NDP52^GE^ variant binds LC3C more efficiently than NDP52^WT^ and this through its cLIR motif. The cLIR-LC3C interaction is crucial for the efficient engulfment of cargoes into the autophagosomes [48], thus the enhanced interaction with LC3C, induced by the presence of the E in position 140 [16] could promote a more efficient autophagic clearance of cargo selected by NDP52 such as pathological hyperphosphorylated TAU proteins (similarly to what has been previously observed for damaged mitochondria [16]). Interestingly, the residues of LC3C crucial for the binding to the cLIR motif of NDP52, are highly conserved among Atg8 orthologs [48,49]. In particular, *Drosophila* Atg8a protein (dAtg8a), homologous to mammalian LC3, conserves all the residues responsible for the preferential binding of hLC3C to hNDP52, with the exception of a single residue (Phe33 in hLC3C replaced by Tyr in dAtg8a) [48,49]. Our findings that, also in *Drosophila melanogaster*, NDP52^GE^ clears pathological phosphorylated TAU more efficiently than NDP52^WT^ might therefore sustain the hypothesis of a pivotal role of the NDP52^GE^/LC3C binding in promoting TAU degradation. However, we cannot exclude that other mechanisms allow NDP52^GE^ to promote autophagy better than NDP52^WT^. While NDP52/LC3C interaction allows cargo targeting to the nascent phagophore, the binding to LC3A/B allows autophagosome maturation, ensuring efficient cargo degradation [14]. Even though the interaction with LC3C was the strongest among the ATG8 family members, we showed that, NDP52^GE^ binds LC3B more efficiently than NDP52^WT^. Thus, it would be possible that also the increased affinity towards LC3B contributes to the ability of NDP52^GE^ to drive autophagy more efficiently than NDP52^WT^.

In addition, it is known that NDP52 oligomerizes and in the context of mitophagy this facilitates the recruitment of the autophagic machinery around mitochondria, for a rapid degradation [50]. We cannot exclude the possibility that the NDP52^GE^ variant is able to oligomerize more efficiently than NDP52^WT^. This would promote a stronger recruitment of the autophagic machinery around the pathological phosphorylated TAU, resulting in a more rapid cargo degradation.

Moreover, it has been shown that NDP52 restricts seeded TAU aggregation by diverting TAU seeds to autophagy, contributing in this way to limit the amplification and propagation of TAU inclusions throughout the brain [51]. The question of whether the NDP52^GE^ is more effective than the NDP52^WT^ needs to be tested. Ultimately, a recent paper by Santos et al. showed that autophagy impacts on NDP52 nuclear localization and links NDP52 nuclear functions to gene transcription and DNA structure regulation [52]. It would be interesting in the future determine whether NDP52 affects TAU degradation also indirectly, by upregulating transcription of factors involved in TAU clearance as well as whether NDP52^GE^ has a greater nuclear localization and/or activity compared to NDP52^WT^.

Lastly, this work introduced a new element that could affect the regulation of NDP52 functions. Indeed, we showed that Protein Phosphatase 2A (PP2A) dephosphorylates NDP52 (both NDP52^WT^ and NDP52^GE^) *in vitro*. This phosphatase is the primary TAU phosphatase in the human brain [32], and plays a crucial role in the AD process [33]. Moreover, in the context of selective autophagy, the cargo receptors require phosphorylation(s) to promote their interaction with ATG8 family proteins [28,29] and the interplay between kinases and phosphatases control these phosphorylation sites in order to modulate the autophagic process [30]. We do not exclude that other phosphatases, as well as PP2A, may contribute to the dephosphorylation of NDP52. In a phospho-proteomic experiment performed by Hoermann et al., a C-terminal peptide of NDP52 was found to be dephosphorylated by both PP2A and PP1 catalytic subunits [53]. In the future, it will be important to study the specific contribution of these and/or other phosphatases to the dephosphorylation of NDP52 in order to better define how this autophagic receptor may be regulated during selective autophagy, in general, and in Alzheimer Disease, in particular.

Of note, NDP52 expression was confirmed in neurons but also in glial cells [15]. Given that microglia-driven neuroinflammation is a key factor that favor TAU pathology development and spreading in AD [51,54,55]. We don’t know if the beneficial role of NDP52^GE^ is limited to neurons or not, and if it also affects neuroinflammation. It should be noted that we originally discovered this variant as a protective factor against Multiple Sclerosis (MS), an autoimmune disease of the central nervous system that is accompanied by neurodegeneration. In this context, we found that NDP52^GE^ was able to eliminate damaged mitochondria more efficiently than NDP52^WT^, and this could in turn reduce the inflammation associated to MS [16]. Mitochondrial dysfunctions have also been shown to be associated with AD [56] thus, in the future, it will be required to better understand whether the beneficial effects of NDP52^GE^ in the context of AD also concern its ability to eliminate damaged mitochondria more efficiently than NDP52^WT^.

Overall, our work highlights the variant NDP52^GE^ as a resilience factor in AD and underlines its robust effectiveness to slow pathological TAU accumulation in *in vitro* and *in vivo*, supporting a great potential for therapeutic interventions in AD patients. Considering the current challenges in finding effective treatments to slow the progression of AD and to improve patients ‘quality of life, this work may serve as a template for future explorations in the field of therapeutic chemistry and translational medicine.

## MATERIALS AND METHODS

### Genetic association case-control analysis

#### Study cohorts

The study cohort overall involved 434 patients affected by sporadic AD and 1000 healthy subjects as control group. In particular, blood samples from 189 AD patients and 1000 control subjects already stored in the Genomic Medicine Laboratory of Santa Lucia Foundation IRCCS were analyzed. AD patients were characterized by a F:M ratio=66:34 and a mean age±s.d.= 74.9 ± 7.55. Briefly, these patients were recruited from 2010 to 2021 at the Outpatient Memory Clinic of the Laboratory of Neuropsychiatry of IRCCS Santa Lucia Foundation, Rome, Italy. The diagnosis of sporadic AD’s dementia was based on medical history and neurological examination, including brain imaging and instrumental tests, overall fulfilling the clinical criteria of the National Institute on Aging and the Alzheimer’s Association [57]. A more detailed description can be found in [58]. These patients were enrolled for a previous study approved by the Ethical committee (CE/PROG.650 approved on 01/03/2018) of IRCCS Santa Lucia Foundation Hospital of Rome and in accordance with the Declaration of Helsinki. A written informed consent of the study was obtained for all patients and control subjects. In addition, blood samples from 245 AD untreated patients were collected from 245 untreated patients admitted to the memory clinic of CRIC in Padua between April 2001 and March 2018. Diagnosis of late onset non-familial AD was made, according to internationally established criteria. The diagnosis was obtained at the end of the diagnostic protocol by clinical, laboratory, and neuropsychological assessment, all subjects underwent CT or MR neuroimaging and 84/245 were further characterized after PET scanning or CSF biomarkers evaluation. All peripheral blood samples were collected after obtaining the written informed consent of the study participants, in accordance with the Helsinki Declaration. The study protocol "Analisi delle Basi Biologiche della Malattia di Alzheimer eredofamiliare e delle forme sporadiche tardive” (3950/AO/2016) was approved by the local ethic committee of the recruitment center on October 06^th^ 2016.

#### Genomic DNA extraction

Genomic DNA was extracted from 200-400 µL of whole blood by means of MagPurix Blood DNA Extraction Kit using MagPurix Automatic Extraction System (Resnova) according to the manufacturer’s instructions. The obtained DNA samples were assessed for quantity and purity byT by DeNovix Spectrophotometer (Resnova), obtaining a concentration range of 50–150 ng/µL and A260/230 and A260/280 ratios included between 1.7 and 1.9.

#### Genotyping analysis

The extracted DNA was employed for the genotyping analysis for the rs550510 (G/A) variant of interest using a predesigned TaqMan assay on QuantStudio® 5 Real-Time PCR System (Applied Biosystems). Each real-time PCR run was performed using a negative control and three positive samples that were previously tested by direct sequencing (BigDye Terminator v3.1) and run on ABI3130xl (Applied Biosystems).

#### Statistical analyses

The genotyping results related to all patients and controls have been tested for evaluating the association and the effect of the rs550510 (G/A) variant as described in [16]. Briefly, as reference group, 1000 healthy subjects were employed. The Hardy–Weinberg equilibrium was confirmed in both cases’ and controls’ cohorts. The obtained data were then evaluated by calculating a P value (*p*) through a 2 × 2 (allele association) and 2 × 3 (genotype association) contingency tables. The statistical associations were considered significant for *p* < 0.05 with a 95% confidence interval. ORs were calculated for evaluating the strength of the association.

### Cell lines, transfection and reagents

SH-SY5Y cells came from an in-house stock and were maintained in DMEM-F12 Glutamax (Gibco, 31331-028), 10% Fetal bovine Serum (FBS) (Corning, 35-015-CV). Human SH-SY5Y P301S-TAU inducible cell line was a kind gift from Luc Buée [34] and were maintained in DMEM complemented with glutamax, non-essential aminoacids, P/S, and 10% of FBS. To induce P301S-TAU expression, cells are maintained in medium with 1ug/ml tetracycline. HeLa cell line harboring endogenously expressed HA-tagged LC3C was a kind gift from C. Behrends [17] and was cultured in DMEM with 10% FBS, 5% glutamine and selected in 4 µg/ml of blasticidin (GIBCO, A1113903). Cells were maintained at 37°C in a humidified atmosphere containing 5% CO_2_ and were trypsinized using 0,05% Trypsin-EDTA (Gibco, 25300-054). Transient DNA transfections were performed using Lipofectamine 2000 according to the supplier’s instructions (Invitrogen, 11668019). To induce hyper-phosphorylation of TAU, cells were treated with Okadaic Acid (OkA) (Santa Cruz Biotechnology, SC-3513). According to supplier’s instructions, OkA was dissolved at 100μM in dimethyl sulphoxide (DMSO) (Sigma-Aldrich, D5879), and used at the final concentration of 100nM for 1h, 1,5h or 2h. An equal amount of DMSO used in the OkA dilution was added to the control cells. To inhibit the autophagic process human SH-SY5Y P301S-TAU inducible cells were treated with 3-Metyladenine (3-MA) for 8h.

### Plasmids

HA-LC3C, FLAG-NDP52^WT^ and FLAG-NDP52^GE^ vectors were previously described [16]. DNA of hNDP52^WT^ or hNDP52^GE^ was subcloned as EcoRI/BamHI fragments into pEGFP-C1 vector for mammalian expression or subcloned into pUAST-attB vector, using EcoRI/BglII restriction sites, to generate *Drosophila* transgenic strains. p3xFLAG-CMV10-NDP52^WT^ or p3xFLAG-CMV10-NDP52^GE^ were used to generate the V136S and τιLIR (from 203-DYWE-206 to 203-AAAA-206) mutant constructs by using the QuickChange Multi Site-Directed Mutagenesis kit (Agilent Technologies, 200515) according to the manufacturer’s instructions. The sequences used for mutagenesis are as follows:

FLAG-NDP52^V136S^: 5’-GGAAGACATCCTGGTTTCTACCACTCAGGGAGAGGTG -3‘; FLAG-NDP52^GE_V136S^: 5’-GGAAGACATCCTGGTTTCTACCACTCAGGAAGAGGTG – 3‘;

FLAG-NDP52^τιLIR^ and FLAG-NDP52^GE_^ ^τιLIR^:

5’-GAAAGAACAGAAGGCCGCTGCGGCGACAGAGCTGCTTCAACTG -3‘.

The constructs encoding GFP-GABARAP, GFP-GABARAPL1 and GFP-GABARAPL2 were kindly provided by Dr P. Grumati (TIGEM, Naples, Italy. The constructs encoding HA-LC3A and HA-LC3B were kindly provided by C. Behrends (Ludwig-Maximilians-Universität (LMU) München, Germany). All constructs were systematically verified by DNA sequencing (BioFab Research, Rome, Italy).

### Western Blotting analysis

Subconfluent cultures of SH-SY5Y cells were lysed in ice-cold lysis buffer (10mM Tris-HCl pH7.4, 150mM NaCl, 1mM EGTA, 0,25%Na-DOC, 0,1% SDS, 1% NP-40) supplemented with 1mM NaF, 1mM Na3VO4, 1mM phenylmethyl sulphonyl fluoride (PMSF) and protease inhibitors (Roche Diagnostic, 11836153001). Samples were kept on ice for 30 minutes, then lysates were clarified by centrifugation at 13000rpm (=15700rcf) for 10 min at 4°C. The concentration of proteins in the supernatants was determined using Bradford assay (Bio-Rad, 5000006). Supernatants were mixed with 4 × Laemmli sample buffer, boiled, resolved by SDS–PAGE and wet transferred over-night to PVDF (Millipore, Immobilon-P IPVH00010). Membranes were blocked with 5% non-fat dry milk in Tris-HCl-buffered saline (TBS) and incubated with the indicated primary and secondary antibodies diluted in 1%milk-TBS1x-0,025% tween-20 and then detected with Clarity Western ECL Substrate (Millipore, Immobilon, WBKLS0500). Densitometry analyses of western blots were performed using the ImageJ software. Antibodies used were as follow: mouse anti-FLAG M2 antibody (Sigma-Aldrich F1804), rabbit anti-FLAG (Sigma-Aldrich F7425), rabbit anti-HA (Sigma-Aldrich, H6908); rabbit anti-β-ACTIN (Sigma Aldrich, A2066); mouse anti-TAU (TAU46) (Santa Cruz Biotechnology, sc-32274); rabbit anti-Phospho-TAU Ser396 (Invitrogen, 44-752G); mouse anti-phospho-TAU Ser202,Thr205 (AT8) (Invitrogen, MN1020); mouse anti-phospho-TAU Thr212,Ser214 (AT100) (Invitrogen, MN1060); mouse anti-VINCULIN (Santa Cruz Biotechnology, sc-73614 clone 7F9); rabbit anti-NDP52 (Cell Signaling, 60732); mouse anti-NDP52 (Novus Biologicals, NBP2-03246, clone OTI4H5); mouse anti-PP2Ac (MERCK Millipore, 05-421 clone1D6); rabbit anti-GABARAP+GABARAPL1+GABARAPL2, used to recognize Drosophila Atg8 (dAtg8) (Abcam; ab109364) ; rabbit anti-GIOTTO (was a gift from GL Cestra; https://www.antibodyregistry.org/AB_2892585); goat anti-mouse IgG (H+L)-HRP conjugated (Bio-Rad, 1706516); goat anti-rabbit IgG (H+L)-HRP conjugated (Bio-Rad, 1706515).

### Co-Immunoprecipitation (Co-IP)

Subconfluent cultures of SH-SY5Y cells were lysed in ice-cold lysis buffer (50mM Tris-HCl pH7.4, 150mM NaCl, Na-DOC 0,25%, Triton 1% NP-40 0,5%, glicerolo 10%) supplemented with 1mM NaF, 1mM Na3VO4, 1mM phenylmethyl sulphonyl fluoride (PMSF) and protease inhibitors (Roche Diagnostic, 11836153001). Samples were kept on ice for 30 minutes, then the lysates were clarified by centrifugation at 13000rpm (=15700rcf) for 10 min at 4°C. 500ug of total proteins from each sample were incubated with mouse anti-HA (Sigma-Aldrich, H3663) or mouse anti-FLAG M2 antibodies (Sigma-Aldrich F1804) on a rotational shaker for 2h at 4 °C, and then incubated with Protein A-Agarose beads (Roche 11134515001) on a rotational shaker at 4 °C for 1 h. Samples were washed with three changes of Washing Buffer (50mM Tris-HCl pH7.4, 150mM NaCl, NP-40 0,5%) supplemented with 1mM NaF, 1mM Na3VO4, 1mM phenylmethyl sulphonyl fluoride (PMSF) and protease inhibitors (Roche Diagnostic, 11836153001). For each change, samples were centrifugated at 2400rpm (=500rcf) for 3 minutes at 4°C, and the supernatants carefully removed. After the final wash, the pellets were mixed with 4×Laemmli sample buffer and western blotting analysis was performed as described above.

### Immunofluorescence

SH-SY5Y cells, grown on coverslips, were washed in warm PBS1x (GIBCO, BE17-512F), and fixed with 4% paraformaldehyde in PBS1x for 10 min at 37°C. After permeabilization with 0.4% Triton X-100 (Sigma-Aldrich, X-100) in PBS 1x for 5 min, cells were incubated at 4°C for 24h in PBS1x, 2% normal goat serum (Sigma-Aldrich, G9023) and the primary antibodies. Cells were then washed with PBS1x and incubated for 1h in PBS1x, 2% normal goat serum and the labeled secondary antibodies. After washes with PBS1x, cells were stained with 1 µg/ml of 4,6-diamidino-2-phenylindole (DAPI) in order to detect the nuclei and coverslips were mounted with Fluoromount Mounting Media (Sigma-Aldrich, F4680). Antibodies used were as follow: mouse anti-HA (Sigma-Aldrich, H3663), rabbit anti-NDP52 (Cell Signaling, 60732), mouse anti-TAU (TAU46) (Santa Cruz Biotechnology, sc-32274), rabbit anti-FLAG (Sigma-Aldrich F7425), mouse anti-FLAG M2 antibody (Sigma-Aldrich F1804), anti-mouse Alexa Fluor^TM^ 488 (Thermo Fisher Scientific, A11017), anti-mouse Alexa Fluor^TM^ 555 (Thermo Fisher Scientific A21425), anti-rabbit Alexa Fluor^TM^ 488 (Thermo Fisher Scientific, A11070) and anti-rabbit Alexa Fluor^TM^ 555 (Thermo Fisher Scientific, A21430). Figure 1C and S2F: samples were analyzed with a Zeiss LSM 800 microscope equipped with 63x oil-immersion objectives. Images were acquired using ZEN system (ZEISS Germany). All acquisitions were performed in non-saturated serial Z-stacks, taking care to cover the entire cell volume. Colocalization analysis in figure 1C was performed by measuring, for each transfected individual cell, the Pearson’s correlation coefficient through the JACoP plugin [59] of the IMAGEJ software [60]. Figure 6: samples were acquired using Nikon Eclipse Ti microscope equipped with 100x oil-immersion objective. TAU intensity was measured using ImageJ/Fiji software [60].

### Lambda PP treatment

SH-SY5Y treated with Okadaic Acid (OkA) as described above were lysed in ice-cold lysis buffer (50mM Tris-HCl pH7.4, 150mM NaCl, Na-DOC 0,25%, Triton 1% NP-40 0,5%, glycerol 10%) supplemented with 1mM PMSF, 2mM DTT and protease inhibitors (Roche Diagnostic, Germany, 11836153001). Samples were kept 15 minutes on ice, then the lysates were clarified by centrifugation at 13000rpm (=15700rcf) for 10 min at 4°C. Supernatants were collected and 10xλPP buffer and 10xMnCl_2_ were added. Each sample was split in two fractions, one was left untreated (as control) while in the other 2μl of λPP enzyme (400U/μl; Santa Cruz sc-200312A) were added. After incubation at 30°C for 1h, samples were mixed with 4×Laemmli sample buffer and western blotting analysis was performed as described above.

### *In vitro* phosphatase assay with purified PP2Ac

Anti-FLAG immunoprecipitation was performed from SH-SY5Y overexpressing FLAG-NDP52^WT^ or FLAG-NDP52^WT^ and treated with OkA 100nM for 1h. After incubation with the beads, samples were washed twice in Washing Buffer and twice in Protein Phosphatase (PP) assay buffer (50mM HEPES pH 7.5; 100mM NaCl; 2mM MgCl2; 1mM DTT; 0.5% NP-40). Samples were then split in two fractions. One fraction was left untreated (as control), and in the other 1μl of purified PP2Ac (0.44mg/ml Cayman Chemical 10011237) was added. After incubation at 30°C for 30min, samples were mixed with 4×Laemmli sample buffer and western blotting analysis was performed as described above.

### *Drosophila melanogaster* strains and procedures

Drosophila stocks were maintained on Drosophila standard medium (Nutri-fly Genesee Scientific, El Cajon, CA, USA) at 25 °C unless otherwise specified. Fly line carrying UAS-hTAU^2N4R^ (51362 (w[1118]: P{w[+mC]=UAS-Tau.wt}1.13); RRID:BDSC_51362) was obtained from the Bloomington Stock Center (https://bdsc.indiana.edu). Fly lines carrying hNDP52^WT^ or hNDP52^GE^ were generated by phiC31-mediated transgenesis by BestGene Inc (**{** HYPERLINK "https://www.thebestgene.com/HomePage.do" **<u>}</u>**). pUAST-attB constructs were inserted on the same genomic landing site on the third chromosome. Eye or panneuronal transgene expression was achieved using the eyeless-GAL4 or elav-GAL4 driver, respectively. <u>Eye area:</u> from adult flies of both sexes were analyzed. Pictures of fly eyes were taken at the higher magnification using the stereomicroscope (ZEISS Semi 508, 50X) (Zeiss) equipped with the Axiocam camera 105 and exploiting ZEISS ZEN software (Blue edition 3.1). Eye area was then measured using NHI ImageJ FIJI freeware software (National Institute of Health, Bethesda, USA) [60]. Measurements were showed using a violin plot in Graphpad Prism 8. More than 150 eyes per genotype were analyzed. For the <u>climbing assay:</u> flies were crossed and maintained at 29°C. Negative geotaxis assays were performed according to the standard protocols [61]. Since the viability of male flies expressing hTAU was strongly reduced in these conditions, we assessed locomotory activity of female adult flies. Female flies were collected and placed in new vials for 24h to eliminate the effect of CO_2_ anesthesia. The flies were then transferred in empty vials for 1h before to be placed in the climbing assay holder. The climbing ability has been scored by tapping flies to the bottom of the vial and scoring how many flies have reached the target line (4 cm) after 10 seconds. Flies were tested in batches of 10-15 and three trials were performed on each analysis. More than 40 flies per genotype were employed in this assay. Data were reported using a Box and whiskers (min to max) plot in Graphpad Prism 8. <u>Lifespan</u> was assessed maintaining groups of 5 virgin flies of same sex in each vial (more of 100 female and 40 males per genotype) and keeping them at 29°C. Flies were transferred on fresh food daily and death events were recorded until the last survivor was dead. Data were plotted on prism GraphPad survival tables. Elapsed days were entered on the X axis and the percentage of survival on the Y axis. Mantel-Cox analysis was used to test the statistical significance of pairs of survival curves. <u>Western blotting analysis</u>: for each genotype, 10 heads of adult flies from both sexes were homogenized. The lysis buffer and the western blotting procedures were performed as described in the previous Western Blotting analysis paragraph.

### *In situ* Proximity Ligation Assay (PLA)

PLA was performed on cells grown on coverslips, fixed in 4% paraformaldehyde and permeabilized with 0.1% Triton X-100-PBS1x for 5minutes. The Duolink *in situ* PLA kit (Sigma-Aldrich DUO92101) was used according to manufacturer’s instructions. The amplification time was 140 min for all tested interactions. Primary antibodies (codes specified in the Western Blotting analysis section) were used in pairs to detect the interactions, or alone as negative control. Figure 1D: HeLa cells harbouring endogenously HA-tagged LC3C were transfected with plasmids coding GFP-NDP52^WT^ or GFP-NDP52^GE^. To detect the LC3C-NDP52^WT^ and LC3C-NDP52^GE^ interactions, mouse anti-HA and rabbit anti-NDP52 primary antibodies were used. Figure S2F: SH-SY5Y were treated with 100nM OkA for 1,5 h at 37 °C. To detect the interactions between endogenous NDP52 and the different hyperphosphorylated forms of TAU, primary antibodies were used as follows: rabbit anti-NDP52 and mouse anti-TAU; rabbit anti-pSer396TAU and mouse anti-NDP52; rabbit anti-NDP52 and mouse anti-AT100. Figure 4: SH-SY5Y were transfected with plasmids coding FLAG-NDP52^WT^ or FLAG-NDP52^GE^ and after 8h of expression treated with DMSO (basal condition) or with 100nM OkA for 1,5 h at 37 °C. To detect the interactions between NDP52 constructs and the different forms of TAU, primary antibodies were used as follows: rabbit anti-FLAG and mouse anti-TAU; rabbit anti-pSer396TAU and mouse anti-FLAG; rabbit anti-FLAG and mouse anti-AT100. Cell nuclei were stained and slides mounted on coverslips using the manufacturer’s mounting medium containing DAPI. Samples were analyzed using the Nikon Eclipse Ti2-E confocal spinning disk inverted microscope. Serial Z stacks were acquired, taking care to cover the entire cell volume. PLA dots were counted using Cell Profiler software [62].

### Correlation analysis

Data of allele or genotype frequencies among countries were obtained from the 1000 Genomes Project Consortium [37]. Incidence and prevalence data of AD and other dementia were obtained from [39]. In particular, we used the Estimated Annual Percentage Change (EAPC) of Age-Standardized Incidence Rate (ASIR) and the EAPC of age-standardized prevalence rate (ASPR) that represent the time trends of age-standardized rates in a specific time period (in our case from 1990 to 2019). These data were produced by Li et al. and were based on the data obtained from 2019 Global Burden Disease (GBD) database, in 21 GBD regions [39,63]. Among these 21 GBD regions, divided on the basis of geographic location, we selected those for which population the 1000 Genomes database reports genetic data. We performed correlation analysis using the resulting 10 GBD region and named them as reported in Li et al and in GBD. Correlation analyses were performed measuring Pearson’s Correlation Coefficient and two-tailed p-value by Prism8 software.

### Statistical Analysis

All statistical calculations were carried out and represented graphically using GraphPad Prism 8 software.

## Data Availability

All data produced in the present study are available upon reasonable request to the authors

## ABBREVIATIONS

AD: Alzheimer Disease;
ATG8: Autophagy-related protein 8;
cLIR: non-canonical LC3 interacting region;
LIR: LC3 interacting region;
NDP52: Nuclear Dot Protein 52, also known as
CALCOCO2: calcium binding and coiled-coil domain 2;
OkA: Okadaic Acid;
PP2A: protein phosphatase 2A;
pTAU: phospho-TAU.

## ACKNOWLEDGMENTS

We thank Dr A. Codemo who performed a part of the clinical work (diagnosis, treatment) and who followed the AD patients from Padova; Dr. F.D. Naso and Dr. F. Liguori for the useful scientific discussions and suggestions. We acknowledge Prof Christian Berhends for the gift of HA-LC3A, HA-LC3B constructs and HeLa cells harboring endogenous HA-tagged LC3C and Dr Luc Buée for the gift of the human SH-SY5Y P301S-TAU inducible cell line. We thank Dr Paolo Grumati for the gift of GFP-GABARAP, GFP-GABARAPL1, GFP-GABARAPL2 plasmids and Dr Francesca Nazio for sharing reagents. We thank prof. Giulia Guarguaglini, Dr. Federica Polverino and the Nikon Reference Centre at the Institute of Molecular Biology and Pathology (CNR) for support with microscopy experiments concerning *in situ* PLA. We thank Dr. Francesca Romana Pellegrini for support with the use of the Cell Profiler Software.

