## Supplementary figures and legends for "A variant of the autophagic receptor NDP52 counteracts phospho-TAU accumulation and emerges as a protective factor for Alzheimer Disease"

**Figure S1**. NDP52$^{GE}$ binds LC3B more efficiently than NDP52$^{WT}$ in a human neuroblastoma cell line. (**A**) Lysates of SH-SY5Y cells expressing the indicated GFP- and FLAG- tagged proteins were immunoprecipitated with anti-FLAG beads. Samples were analyzed by Western blot using the indicated antibodies. The graph reports the amount of the indicated GFP-GABARAP protein coprecipitated by the corresponding FLAG-NDP52 protein. Data were expressed as percentage variation over FLAG-NDP52$^{WT}$. Images and data are representative of three independent experiments. Data are presented as means ± SEM. (two tailed unpaired *t*-test: not statistical) (**B**) Lysates of SH-SY5Y cells expressing the indicated HA- and FLAG- tagged proteins were immunoprecipitated with anti-HA beads. Samples were analyzed by Western blot using the indicated antibodies. The graph reports the amount of FLAG-NDP52 coprecipitated by the corresponding HA-LC3 protein. Data were expressed as percentage variation over FLAG-NDP52$^{WT}$. Images and data are representative of three independent experiments. Data are presented as means ± SEM. *p<0,05 (two tailed unpaired *t*-test).

**Figure S2.** Okadaic Acid (Oka) treatment in SH-SY5Y cells induces the accumulation of phosphorylated forms of TAU. (**A**) Lysates of SH-SY5Y cells treated with 100nM OkA for the indicated times. Western blot was performed using the indicated antibodies. Images are representative of eight independent experiments. (**B**) Lysates of SH-SY5Y cells treated with 100nM OkA for the indicated times were split and incubated with (+) or without (-) Lambda Protein Phosphatase (λPP). Samples were analyzed by western blot using the indicated antibodies. Images are representative of three independent experiments. The lines in the TAU panel indicate non-phosphorylated (lilac) and phosphorylated (purple) TAU. Arrow heads in the pSer396-TAU panel indicate monomeric (pink) and high molecular weight oligomeric (brown) phospho-Ser396-TAU that disappeared in λPP treated samples and that were measured and summed for the western blot densitometry analyses. (**C**) TAU- and pSer396TAU-specific bands were measured, summed and normalized to the corresponding signal of VINCULIN, used as loading control. Results are expressed as percentage variation of the levels at time 0. Data are representative of eight independent experiments. (**D and E**) Lysates of SH-SY5Y cells treated with 100nM OkA for the indicated times. Western blot analysis was performed using the indicated antibodies. Images are representative of four independent experiments. Arrow head indicates antibody-specific bands that were measured and summed (in the case of AT8) for densitometry analysis. Signals from the indicated TAU antibodies were normalized to the corresponding signal of VINCULIN, used as loading control. Results are shown in the graph on the side of each western blot. Data information: In (C-E) data are presented as means ± SEM. * indicates comparison to 0: * p<0,05; *** p<0,001; ****p = <0,0001. & indicates comparison to 1h: & pvalue<0,05; &&& pvalue < 0,001 (two tailed unpaired *t*-test). (**F**) Representative confocal images of *in situ* PLA performed in SH-SY5Y treated with 100nM OkA for 1,5 h at 37 °C. *In situ* PLA experiments were performed using anti-NDP52/anti-TAU, anti-pSer396TAU/anti-NDP52, anti-NDP52/anti-AT100 primary antibodies. Scale bar: 10μm.

**Figure S3.** PP2A dephosphorylates both $NDP52^{WT}$ and the natural variant $NDP52^{GE}$. *In vitro* dephosphorylation assay. Lysates of SH-SY5Y cells expressing the indicated FLAG-tagged proteins and treated with (+) or without (-) 100nM OkA for 1h were immunoprecipitated with anti-FLAG beads. Immunoprecipitated samples were split and incubated with (+) or without (-) recombinant purified PP2A catalytic subunits (PP2Ac). Obtained samples were analyzed by western blot using the indicated antibodies. FLAG- $NDP52^{WT}$ immunoprecipitated from non-treated SH-SY5Y cells and not incubated with PP2Ac (first lane) was used as unphosphorylated NDP52 standard. IgG are shown as loading control. Images are representative of three independent experiments.

**Figure S4.** hNDP52 partially rescues hTAU phenotypes in *Drosophila melanogaster*. (**A**) Lysates from heads of flies expressing the indicated human (h) transgenes at 25°C under control of eyeless-GAL4 driver (Ey>) were analyzed by western blot using the indicated antibodies. + indicates non-transgenic flies used as control. Images are representative of 5 independent experiments. Signals from the indicated TAU and NDP52 antibodies were measured and normalized to the corresponding signal of GIOTTO, used as loading control. Results, expressed as means ± SEM, are shown in the graphs. # indicates comparison to hTAU: # pvalue < 0,05, ## pvalue < 0,01 (Ordinary One-way ANOVA, Turkey's multiple comparisons test). (**B**) Survival curve showing lifespan of male flies expressing panneuronally (elav-GAL4, ELAV>) the indicated transgenes at 29°C. * indicates comparison to control (+): **** pvalue <0,0001; # indicates comparison to hTAU: # pvalue < 0,05; ## pvalue <0,01 (Log-rank (Mantel-Cox) test). (**C and D**) Lysates from heads of flies expressing the indicated transgenes under control of GMR-GAL4 driver (Ey>) (**C**) or panneuronally (elav-GAL4, ELAV>) (**D**) were analyzed by western blot using the indicated antibodies. The "+" sign refers to non-transgenic flies used as control. Images are representative of at least 4 independent experiments. Signals from NDP52 antibody were measured and normalized to the corresponding signal of GIOTTO, used as loading control. Results, expressed as means ± SEM, are shown in the graphs below (two tailed unpaired *t*-test: not statistical).

**Figure S5.** Correlation analysis between worldwide incidence or prevalence of AD and other dementia and the frequency of the allele G or the genotype GG or the genotype AG. Graphs reporting the correlation analysis between worldwide incidence or prevalence of AD and other dementia and the frequency of allele G (coding for $NDP52^{WT}$) (**A**) or genotype GG (**B**) or genotype GA (**C**). Incidence and prevalence data are the estimated annual percentage changes of age-standardized rates of incidence and prevalence from 1990 to 2019 reported in [1]. Data of allele or genotype frequencies are from the 1000 Genomes Project Consortium [2]. Pearson's Correlation Coefficient (r) and 95%

confidence interval are shown. * pvalue (two-tailed) < 0,05; ** pvalue (two-tailed) <0,005. Correlation analyses were performed with Prism8 software.

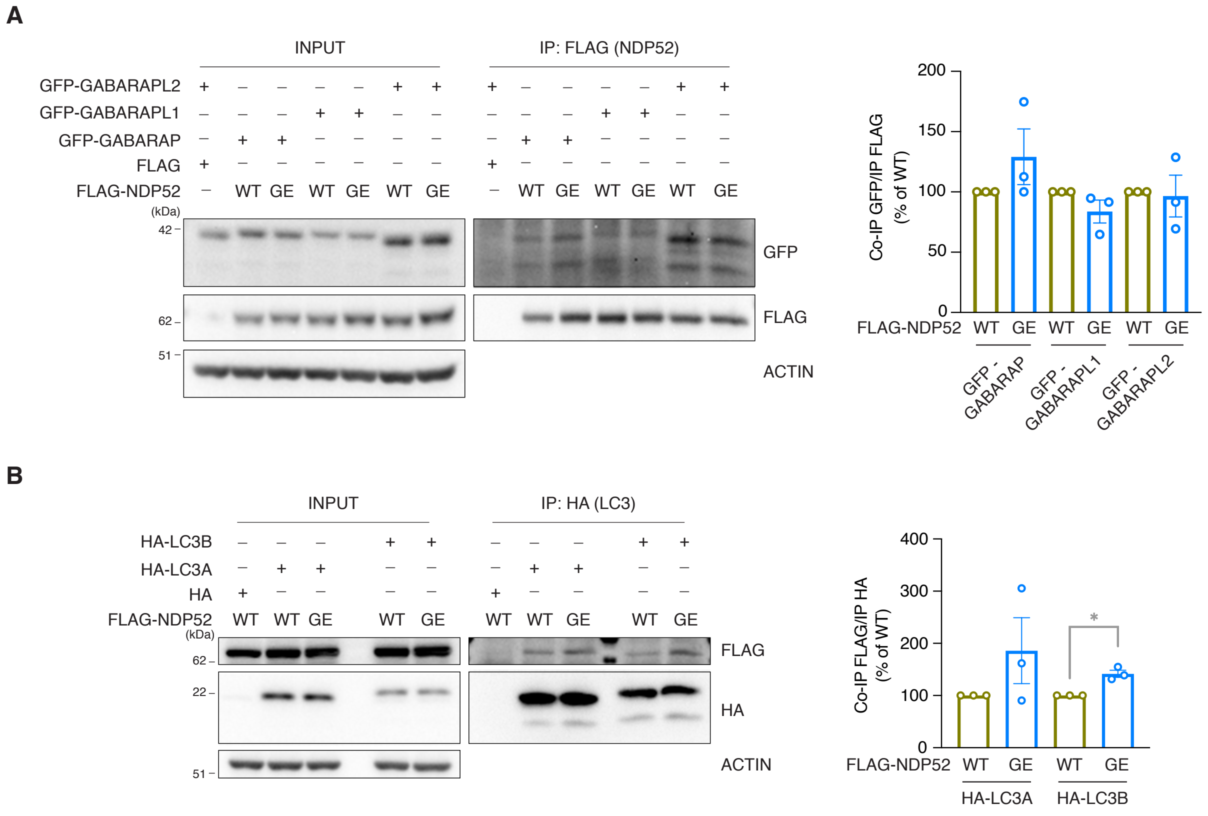
A
INPUT
IP: FLAG (NDP52)
GFP-GABARAPL2
GFP-GABARAPL1
GFP-GABARAP
FLAG
FLAG-NDP52
WT GE WT GE WT GE
(kDa)
42
62
51
GFP
FLAG
ACTIN
Co-IP GFP/IP FLAG (% of WT)
200
150
100
50
0
FLAG-NDP52 WT GE WT GE WT GE
GFP - GABARAP
GFP - GABARAPL1
GFP - GABARAPL2
B
INPUT
IP: HA (LC3)
HA-LC3B
HA-LC3A
HA
FLAG-NDP52
WT WT GE WT GE
(kDa)
62
22
51
FLAG
HA
ACTIN
Co-IP FLAG/IP HA (% of WT)
400
300
200
100
0
*
FLAG-NDP52 WT GE WT GE
HA-LC3A
HA-LC3B

Figure S1

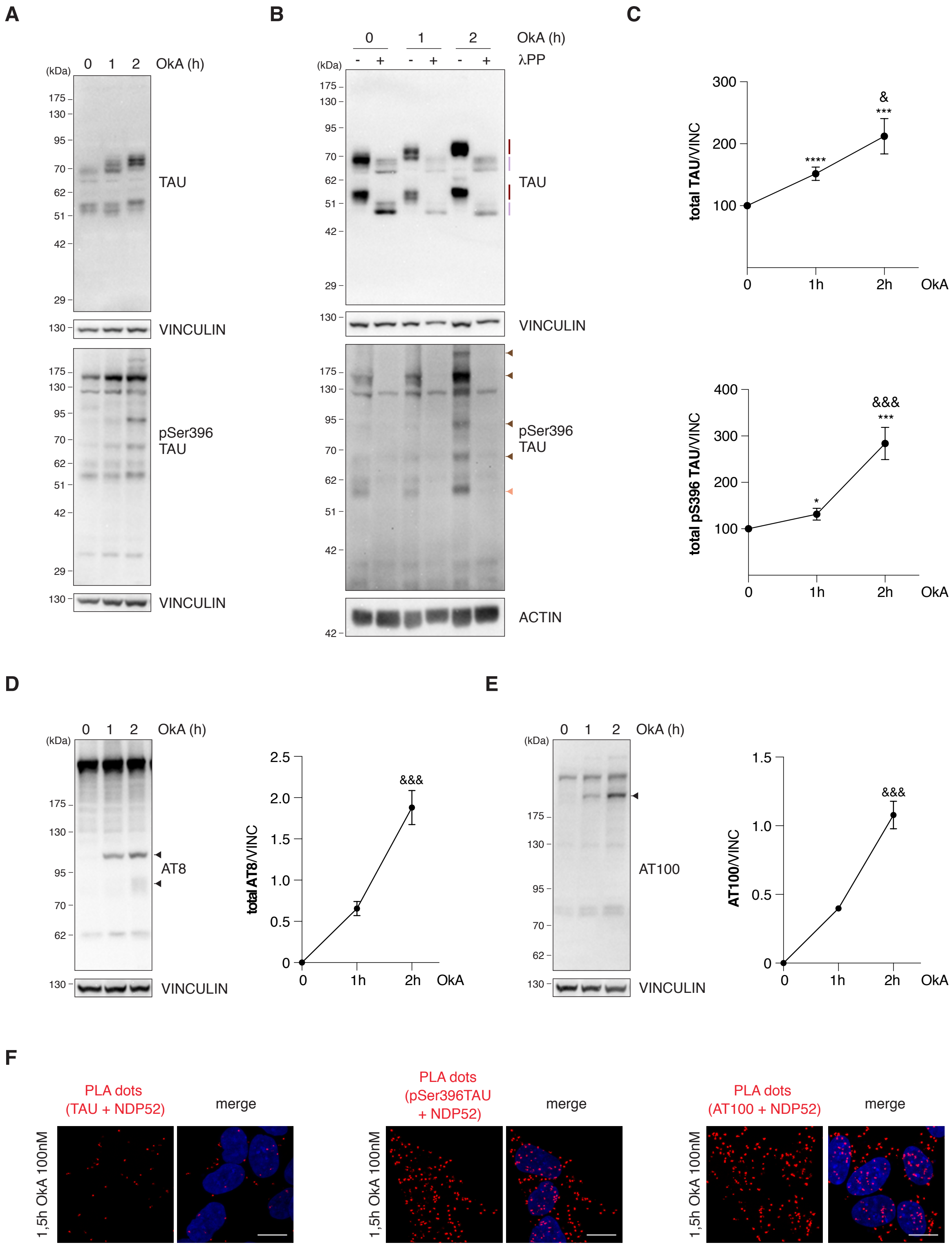

A
B
C
D
E
F
(kDa)
0 1 2 OkA (h)
175
130
95
70
62
51
42
29
TAU
VINCULIN
pSer396
TAU
0 1 2 OkA (h)
- + - + - + λPP
ACTIN
total TAU/VINC
300
200
100
0 1h 2h OkA
&
***
****
total pS396 TAU/VINC
400
&&&
*
AT8
total AT8/VINC
2.5
2.0
1.5
1.0
0.5
0
AT100
AT100/VINC
PLA dots
(TAU + NDP52)
merge
(pSer396TAU
+ NDP52)
(AT100 + NDP52)
1,5h OkA 100nM


Figure S2

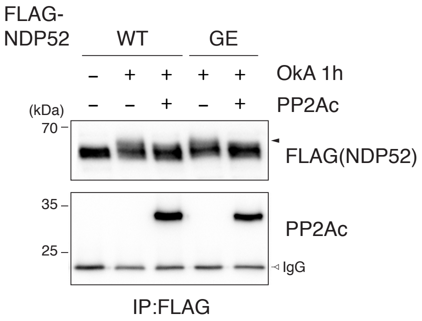
FLAG-
NDP52
WT
GE
– + + + +
OkA 1h
– – + – +
PP2Ac
(kDa)
70
FLAG(NDP52)
35
25
PP2Ac
IgG
IP:FLAG

Figure S3

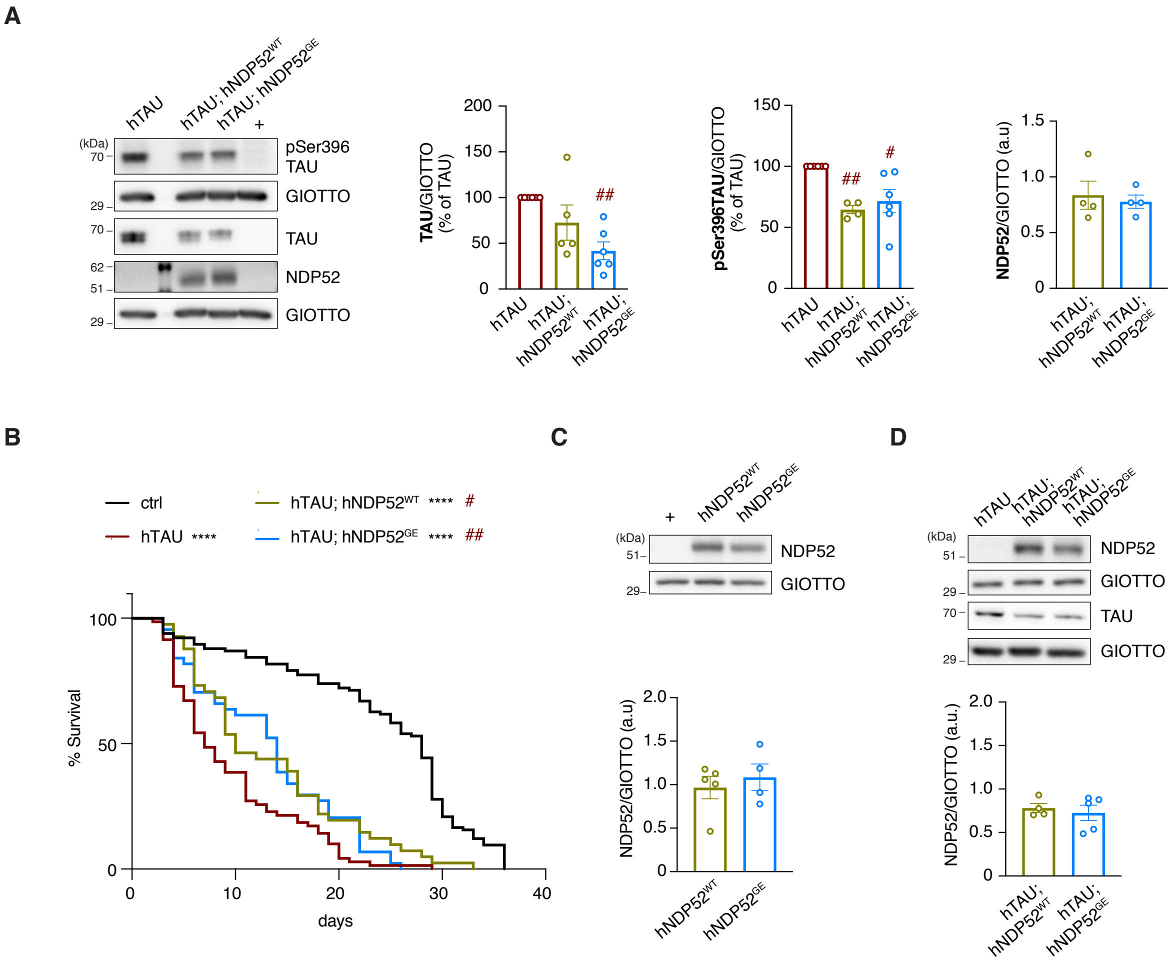
A
hTAU
hTAU; hNDP52WT
hTAU; hNDP52GE
+
(kDa)
70
29
70
62
51
29
pSer396 TAU
GIOTTO
TAU
NDP52
GIOTTO
TAU/GIOTTO (% of TAU)
200
150
100
50
0
##
pSer396TAU/GIOTTO (% of TAU)
150
100
50
0
##
#
NDP52/GIOTTO (a.u)
1.5
1.0
0.5
0
hTAU; hNDP52WT
hTAU; hNDP52GE
B
ctrl
hTAU ****
hTAU; hNDP52WT **** #
hTAU; hNDP52GE **** ##
% Survival
100
50
0
0
10
20
30
40
days
C
+
hNDP52WT
hNDP52GE
(kDa)
51
29
NDP52
GIOTTO
NDP52/GIOTTO (a.u)
2.0
1.5
1.0
0.5
0
D
hTAU
hTAU; hNDP52WT
hTAU; hNDP52GE
(kDa)
51
29
70
29
NDP52
GIOTTO
TAU
GIOTTO
NDP52/GIOTTO (a.u)
2.0
1.5
1.0
0.5
0

Figure S4

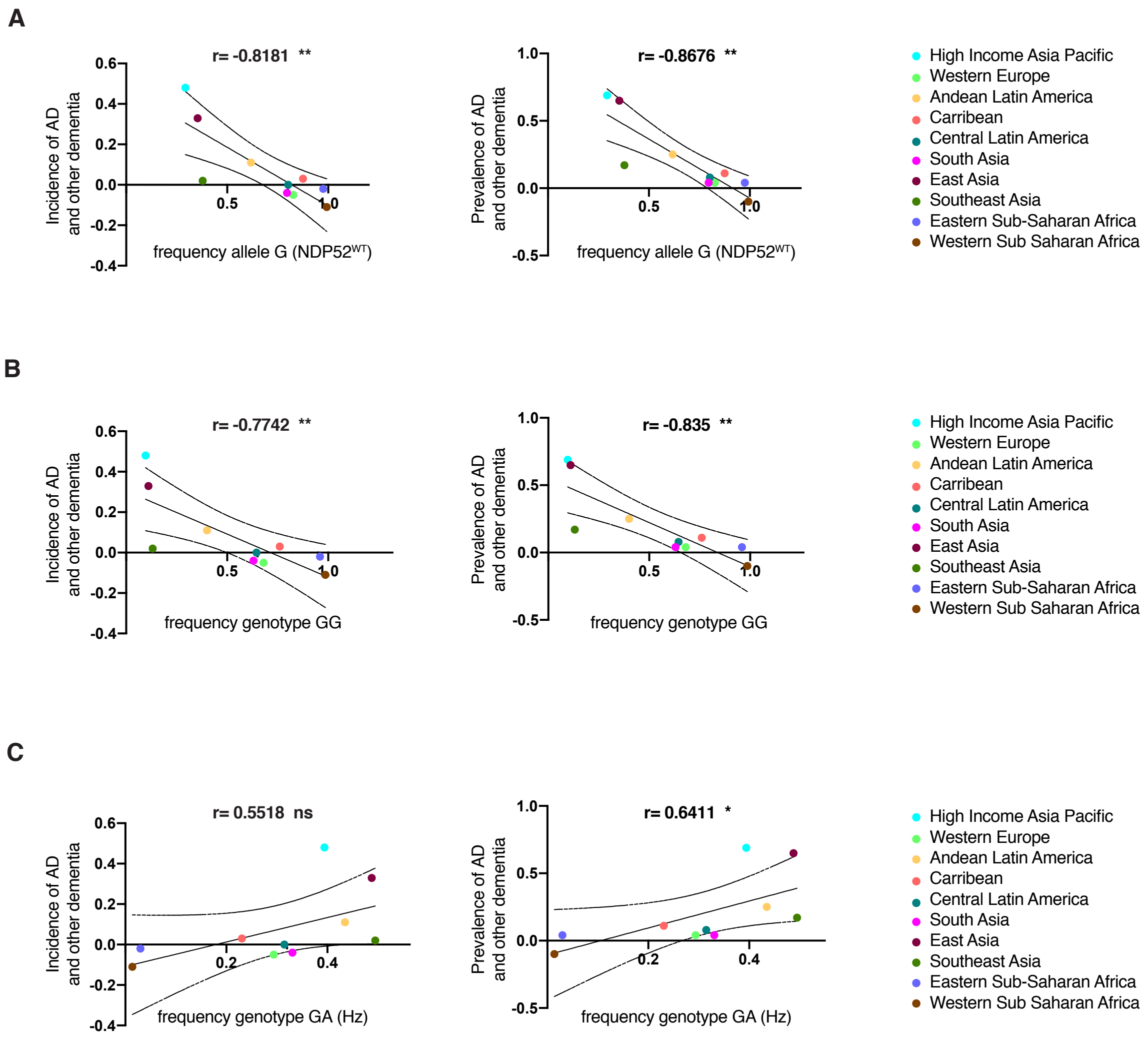

A
r= -0.8181 **
Incidence of AD and other dementia
frequency allele G (NDP52WT)
r= -0.8676 **
Prevalence of AD and other dementia
frequency allele G (NDP52WT)
High Income Asia Pacific
Western Europe
Andean Latin America
Carribean
Central Latin America
South Asia
East Asia
Southeast Asia
Eastern Sub-Saharan Africa
Western Sub Saharan Africa
B
r= -0.7742 **
Incidence of AD and other dementia
frequency genotype GG
r= -0.835 **
Prevalence of AD and other dementia
frequency genotype GG
C
r= 0.5518 ns
Incidence of AD and other dementia
frequency genotype GA (Hz)
r= 0.6411 *
Prevalence of AD and other dementia
frequency genotype GA (Hz)


Figure S5
